# COVID-19 amplified racial disparities in the U.S. criminal legal system

**DOI:** 10.1101/2021.12.14.21267199

**Authors:** Brennan Klein, C. Brandon Ogbunugafor, Benjamin J. Schafer, Zarana Bhadricha, Preeti Kori, Jim Sheldon, Nitish Kaza, Arush Sharma, Emily A. Wang, Tina Eliassi-Rad, Samuel V. Scarpino, Elizabeth Hinton

## Abstract

The criminal legal system in the United States drives an incarceration rate that is the highest on the planet, with disparities by class and race among its signature features [1–3]. During the first year of the COVID-19 pandemic, the number of incarcerated people in the U.S. decreased by at least 17%—the largest, fastest reduction in prison population in American history [4]. In this study, we ask how this reduction influenced the racial composition of U.S. prisons, and consider possible mechanisms for these dynamics. Using an original dataset curated from public sources on prison demographics across all 50 states and the District of Columbia, we show that incarcerated white people benefited disproportionately from the decrease in the U.S. prison population, and that the fraction of incarcerated Black and Latino people sharply increased. This pattern of increased racial disparity exists across prison systems in nearly every state and reverses a decades-long trend before 2020 and the onset of COVID-19, when the proportion of incarcerated white people was increasing amid declining numbers of incarcerated Black people [5]. Although a variety of factors underlie these trends, we find that racial inequities in average sentence length are a major contributor. Ultimately, this study reveals how disruptions caused by COVID-19 exacerbated racial inequalities in the criminal legal system, and highlights key forces that sustain mass incarceration.

## 1 Introduction

Mass incarceration in the United States is distinguished by striking racial disparities and a rate of imprisonment that surpasses all other nations, with 2.12 million people behind bars in 2019 [1–3, 6–9]. Due to a combination of structural inequities and discriminatory enforcement, Black and Latino people are more likely to be stopped by police [10], held in jail pretrial [11], charged with more serious crimes [12], and sentenced more harshly than white people [13, 14]. These practices have made Black men in the U.S. six times as likely and Latino men 2.5 times as likely to be incarcerated as white men [15, 16].

In this study, we demonstrate how the COVID-19 pandemic—which produced the largest, most rapid single-year decrease in prison population in U.S. history—amplified existing inequities in the nation’s criminal legal system [4]. Across nearly every state and federal prison system, we observe a convergent pattern: a substantial decrease in the overall number of people incarcerated (by approximately 200,000), but a significant increase in the proportion of incarcerated Black, Latino, and other non-white people. We conclude that sentencing patterns are a central mechanism driving the racial disparity.

The trend we identify represents a significant deviation from patterns preceding the pandemic. Prior to COVID-19, incarcerated Black people accounted for a declining share of the total prison population: roughly 41.6% of people incarcerated in state prisons were Black in March 2013, and by March 2020 this number had fallen to 38.9%—a decline of 2.7 percentage points over seven years. During the height of COVID-19 closures, from March 2020 to November 2020, this percentage increased by 0.9 points, erasing much of the progress over the last decade (Figure 1B, 2, and A.2; see Figure A.16 for comparison between effects among non-white vs. Black populations). The trend we observe at the national level is reproduced exactly among states with the highest Black and Latino populations, and persists in some form in nearly every other state.

**Figure 1:**
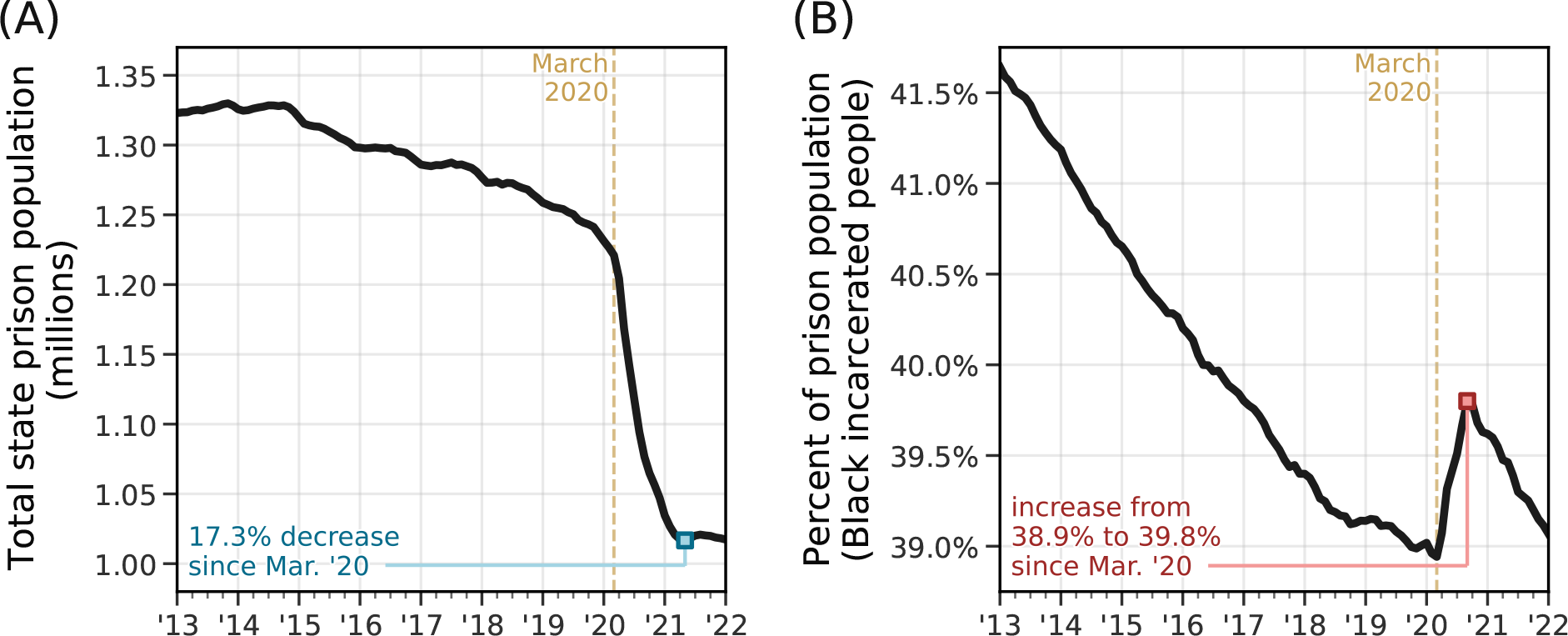
Dynamics of the U.S. prison population. (A) Total number of incarcerated people in the United States from January 2013 to January 2022. **(B)** Total percent of incar-cerated Black people, as reported by states’ Departments of Correction. According to data from the United States census, Black people account for 13.4% of the total population [18]. This plot includes data from 49 states and the District of Columbia—data from Michigan are excluded as the state reports only “white” and “nonwhite” as race categories.

**Figure 2:**
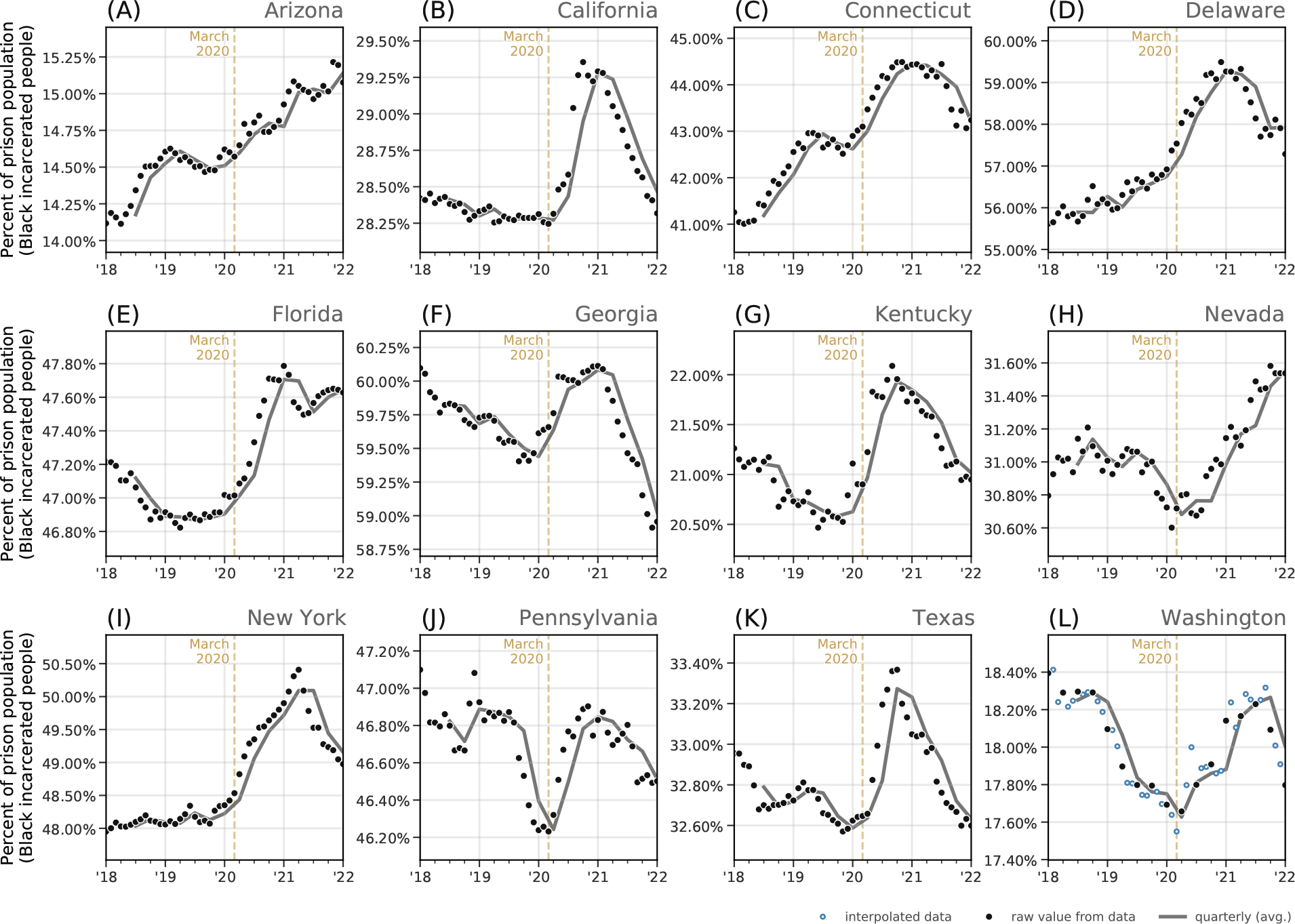
Percent of incarcerated Black people across 12 representative states. In some states, the percent of incarcerated Black people had been decreasing over the last several years. In others, this percentage had been increasing. Across a variety of pre-2020 trends, we see the same general trend across the United States: during the pandemic, incarcerated Black people accounted for an even larger share of the total prison population than in previous years. We plot this trend in **(A)** Arizona, **(B)** California, **(C)** Connecticut, **(D)** Delaware, **(E)** Florida, **(F)** Georgia, **(G)** Kentucky, **(H)** Nevada, **(I)** New York, **(J)** Pennsylvania, Texas, and **(L)** Washington—states with consistent, frequent data reporting.

Data reporting methods on racial demographics in prisons have made it difficult for researchers to disentangle the various mechanisms driving observed disparities in incarcerated populations. We manually assembled and validated a dataset covering all 50 U.S. states, the District of Columbia, and the Federal Bureau of Prisons (FBOP) to both quantify the widening racial disparity observed during the first year of the COVID-19 pandemic and uncover its plausible causes. The result of this newly assembled, public dataset—comprising over 9,000 records across more than 20 years—is an unprecedented view into the dynamics of prison populations before, during, and after the onset of the pandemic.

Overall, the number of incarcerated people decreased dramatically in 2020. But we show that the magnitude of these declines were not equal by race, especially for incarcerated Black people. We estimate that nearly 15,000 fewer Black people would have been incarcerated in January 2021 if the racial disparities we observe were not present (see Table A.6). We discuss this observed disparity and related observations in light of the ethics of public health interventions, national debates about the future direction of policing and incarceration, and the importance of data infrastructure in responsible public policy. These discussions highlight how sentencing and other policies that appear to be “race-blind” can nonetheless lead to outcomes that are skewed by race [17]. We speculate that our findings transcend the influence of COVID-19, and discuss how large-scale disruptions can have a clear, quantifiable signature on extant inequalities.

## 2 Results

### 2.1 Declining incarcerated populations

The population of people incarcerated in U.S. state prisons decreased by at least 17% between March 2020 and July 2021, from approximately 1.23 million to 1.02 million (Figure 1A). A decrease in prison population occurred in every state, and, in most, started in early to mid-April 2020 (see Figure A.1 for a state-by-state look at prison populations over time). This nation-wide trend persisted despite stark differences in state-level trends pre-2020. For instance, some states entered 2020 with a steadily-declining prison population (e.g. Massachusetts, South Carolina, California, among others), others had relatively stable prison populations (e.g. Virginia, Georgia, Iowa, etc.), and many had growing prison population before COVID-10 (e.g. Alabama, Indiana, Montana, etc.) (see Figure A.1). Nevertheless, we see large reductions in the prison population across every state in the U.S. during the pandemic.

As of January 2022, several states’ prison populations continued to decrease (Arizona, Massachusetts, Washington, Louisiana, Pennsylvania, among others) and did so steadily throughout the pandemic. Other states’ prison populations dropped sharply in the early months of the pandemic but saw their prison populations begin to approach pre-pandemic levels by 2022 (North Dakota, South Dakota, Montana, Iowa, among others). In Figure A.1, we plot time series for each state’s prison population over the last several years. Additionally, in Table A.1, we give an overview for each state’s approach for reporting prison population statistics, along with how we collected each state’s data. In Table A.2, we detail the scale and timing of each state’s population decline.

### 2.2 Changing racial demographics in prisons

Despite an overall decline in the total incarcerated population during the pandemic, there was an increase in the proportion of incarcerated Black people (Figure 1B). This increase in racial disparity occurred nationally and in nearly every state, transcending vast differences in approach to crime and incarceration. In Figure 2, we show the percentage of incarcerated Black people across 12 states (see Figure A.2 for these trends in every state and the Federal Bureau of Prisons). However, the spike in the proportion of Black people in prison was temporary in most states, eventually returning to pre-pandemic levels. We explore possible explanations for this reversal in subsequent sections, but the most likely reason is that the pace of prison admissions—which typically have a lower Black-white racial disparity than the overall incarcerated population [19]—began to approach pre-pandemic rates in early 2021.

While the national trend we identify in Figure 1, i.e., an abrupt increase in the proportion of Black people incarcerated, occurred in most state-level prison systems, there were meaningful differences that suggest possible mechanisms behind the disparity (see Figures 2, A.2, A.4, and A.3). In Figure 2, we highlight several examples of state-level variability in the proportion of Black people incarcerated; for instance, states like Georgia, Kentucky, and Texas resemble the shape seen nationally, whereas states like Connecticut and Delaware saw an already-increasing trend in the percent of incarcerated Black people increase even faster after March 2020. Five states—Maine, Maryland, Missouri, Oregon, and Wyoming— are the only prison systems in the U.S. that do not clearly conform to the pattern we see across the country (a few states in Fig. A.2—e.g. Missouri and Oklahoma—technically fit our criteria for exhibiting this trend but only weakly). These five states have a combined incarcerated population that amounts to roughly 5% of the national total and offer important insights into the underlying mechanisms behind the trends we see nationally. Namely, each of these states either has 1) a relatively small proportion of incarcerated Black people or 2) a prison system with fewer people with shorter-term (e.g. fewer than 2 years) sentences compared to nationwide averages. We will show that the latter is likely the more powerful force contributing to the overall nationwide trend.

Ultimately, these observations lead us to outline three explanations that could bring about the trends from Figure 1: 1) who is admitted to prison, 2) who is released from prison, and 3) who remains in prison. These proposed mechanisms demonstrate different levers through which the pandemic may have influenced the racial composition of the incarcerated population, and dovetail with existing research on the dynamics of the American carceral state.

### 2.3 Mechanisms of disparity

Consider a time series of a state’s prison population that does not notably change over several years. In order for this to occur, there needs to be approximately the same number of admissions and releases. In order for the demographic makeup of the prison population to remain stable, the relative number of admissions and releases by race also needs to be roughly equivalent over time. If there are sustained periods with more admissions (or releases) of a certain demographic, that will skew the overall distribution of the prison population.

Understanding the dynamics of admissions, releases, and sentencing offers us a path toward identifying and isolating potential mechanisms that could bring about a steadily declining rate of Black incarceration (seen for nearly a decade prior to the pandemic), and the subsequent spike in the proportion of incarcerated Black people during the COVID-19 pandemic. Namely, the observed spike in Figure 1B must be due to a disparity in who was admitted to prison during the pandemic, who was released, or a combination of both.

### Admissions: Disruptions in court operations

In every state except Nebraska, courts closed at the beginning of the pandemic. These closures substantially reduced or altogether halted admissions into prisons for several months, starting around April 2020 [20–23]. The Virginia Department of Corrections acknowledges the causal effect of court closures on the state’s incarcerated population in their 2020 Annual Report [24], “The reduction in [average daily population] is directly attributed to the suspension of intake due to COVID-19.” Similarly, a spokesperson for the Michigan Department of Corrections estimated that half of the reductions in incarcerated population were due to a decline in new admissions from courts and county jails [25].

Based on admissions data from 18 states, we estimate that the total monthly admissions to prison fell to about 30% of pre-pandemic averages by May/June 2020 (see Figure 4). This reduction in admissions provides a potential mechanism behind the sharp increase in the percent of incarcerated Black people in Figure 1B. Specifically, systematic racial differences occurring in monthly prison admissions during this period could drive changing disparity in the demographics of the incarcerated population. However, data from the 18 states presented in Figure 4C actually show the reverse, i.e., the percent of Black individuals admitted to prison fell even lower than the corresponding rate for white admissions. While we do see abrupt spikes in the percent of Black individuals admitted to prison in a few states (e.g. Wisconsin, Texas), this proposed mechanism appears not to be widespread enough to explain the nationwide trends we observe.

Data from Florida offers another example of how changes to court proceedings influence prison admissions and the racial distribution of people admitted to prison. In Figure A.18 we plot monthly trial statistics from circuit criminal defendants in Florida; after March 2020, we see sharp declines in the number of disposed defendants in Florida Circuit Criminal Courts, as well as the percentage of filed defendants that become disposed. Amid these declines, we see an abrupt increase in the percentage of cases that were dismissed before trial (i.e., defendants whose charges were dropped). In Figure A.19, we report that an increased proportion of the defendants with pretrial case dismissals were white in the months after the start of the pandemic.

Prison admissions may also decline due to policy changes or disruptions to a common source of prison admissions: county jails. While there continues to be poor standards for reporting and maintaining these kinds of data, this potential source of prison admissions is important for a nationwide story of mass incarceration during the COVID-19 pandemic. Despite variability in admissions playing a role in the racial distribution of incarcerated populations, changing disparity in admissions alone does not appear to be widespread enough to account for the nationwide trends we observe in Figure 1B. As we will see in the following section, a similar story emerges when looking at the demographic of people released from prison.

### Releases: Typical and pandemic decarceration

In an effort to reduce the risk of SARS-CoV-2 transmission, several states enacted policies designed to de-densify prisons. Depending on the state, these directives came from executive orders from the governor, state legislatures, or governing boards. In Utah, for example, policies around releases are designed, approved, and implemented by the Board of Pardons and Parole (BPP)—an entirely separate entity from the courts and the Department of Corrections. According to the BPP, incarcerated people who are eligible for early release needed to be already characterized as a non-violent offender, be within 90 days of release (this was later extended to 180 days [26]), and have an approved address to stay at after their release.

In Arkansas, an authorization from Governor Hutchinson (Executive Orders 20-06 and 20-16 [27]), made 1,243 incarcerated people eligible for early release as of April 30, 2020. Those deemed eligible needed to have a parole plan in place, be medically screened (i.e., tested and screened for symptoms of COVID-19), and undergo final approval by the Arkansas Department of Corrections director in order to be released. In Section A.4.3 and Figure A.23, we show that disproportionately more white people were released in Arkansas through this effort. The racial disparity in who was released by Governor Hutchinson’s orders is due to the overlap between the state’s release eligibility criteria and the racial differences in sentence classification—a tension we discuss further in Section 4.2.2.

In Figure 4D, we plot estimates of the nationwide change in monthly releases as a percent of pre-pandemic values. What we see is that, despite efforts to reduce prison density through targeted releases, the rate of prisoner release was *lower* during much of the pandemic. At its lowest value (between February 2021 - May 2021), the number of people released from prison each month reached nearly 70% of pre-pandemic values. In the absence of changing admission patterns, this decline in releases should have led to an *increase* in the total incarcerated population in the U.S., which is the opposite of the pattern we see in 1A. Therefore, we can conclude rather strongly that changing release rates did not drive the reduction in the incarcerated population during the pandemic. We also do not find meaningful differences in the relative number of releases by race during this time period; if anything, these data suggest that during the early months of the COVID-19 pandemic, Black people accounted for a higher percent of monthly releases compared to pre-pandemic averages. As was the case for demographics of prison admissions, disparities in the monthly releases are unlikely to be driving the trends in Figure 1B. However, data on prison releases points to an important, related process underlying the demographic patterns in incarcerated populations; in the next section, we focus less on those released from prison in any given month, but rather, those who remain.

### Sentencing: Long-standing differences in length of incarceration

Based on demographic data from 18 state prison systems, racial disparities in admissions and releases alone are not able to explain the broad trends observed nationally (see Figure 4). In fact, if these were the only factors influencing prison population demographics, we would expect the opposite effect seen in Figure 1B, since we observe a large increase in the proportion of white admissions after the start of the pandemic, amid large decreases in the proportion of Black admissions and relatively commensurate rates of releases. There are examples of individual states that show sudden increases in the relative amount of Black people admitted to prison at the start of the pandemic (see Texas, for example, in Figure A.20). Similarly, there are examples of large-scale releases causing an abrupt increase in the percent of incarcerated Black people (see a recent example in January 2022 in data from the Federal Bureau of Prisons, Figure A.9). Nevertheless, we do not see these factors as being anywhere near as influential as disparities in sentencing of people already incarcerated at the start of the pandemic.

In short, the most important factor underlying the dynamics in Figure 1B is related to differences in the average sentence length of incarcerated people by race. As a statistical observation this point is quite simple: provided there are (i) differences in the average length of prison sentence by race (e.g. the average incarcerated Black person serving a longer prison sentence than the average white incarcerated person; see Texas as an example in Figure A.22) and (ii) sustained reductions in new admissions (as in Fig. 4C), then we will expect to see the effect observed in Figure 1B. In addition to that basic mechanism, one can imagine factors that would exacerbate and/or attenuate the size and timing of the spike. These include: new or atypical patterns in prison admissions by race (relative to averages prior to the decline in admissions), or new or atypical changes in prison releases by race.

By casting sentencing differences as the driver behind the observations in this study, we are able to better understand why the main effect in Figure 1B is so pronounced among incarcerated Black people and less so (though still present) when looking at incarcerated

Latino people (see Figure 3). Illinois and Texas offer two particularly powerful examples that show increases in their proportion of incarcerated Black people. As one would expect given our proposed mechanism, the median sentence length for incarcerated Black people in each of these states is higher than that of white people. However, when we compare the median sentence lengths between incarcerated Black and Latino people, we find high Black-Latino overlap in Illinois but high white-Latino overlap in Texas. That is, white people in Illinois serve shorter sentences on average than Black and Latino people, but in Texas, white and Latino people serve shorter sentences than incarcerated Black people. According to the mechanism proposed above, we would expect this baseline difference in sentencing lengths to produce pandemic-related spikes in the percent Black and Latino people in Illinois and only produce spikes in the percent incarcerated Black people in Texas. This, in fact, is what we observe (see Figure A.7).

**Figure 3:**
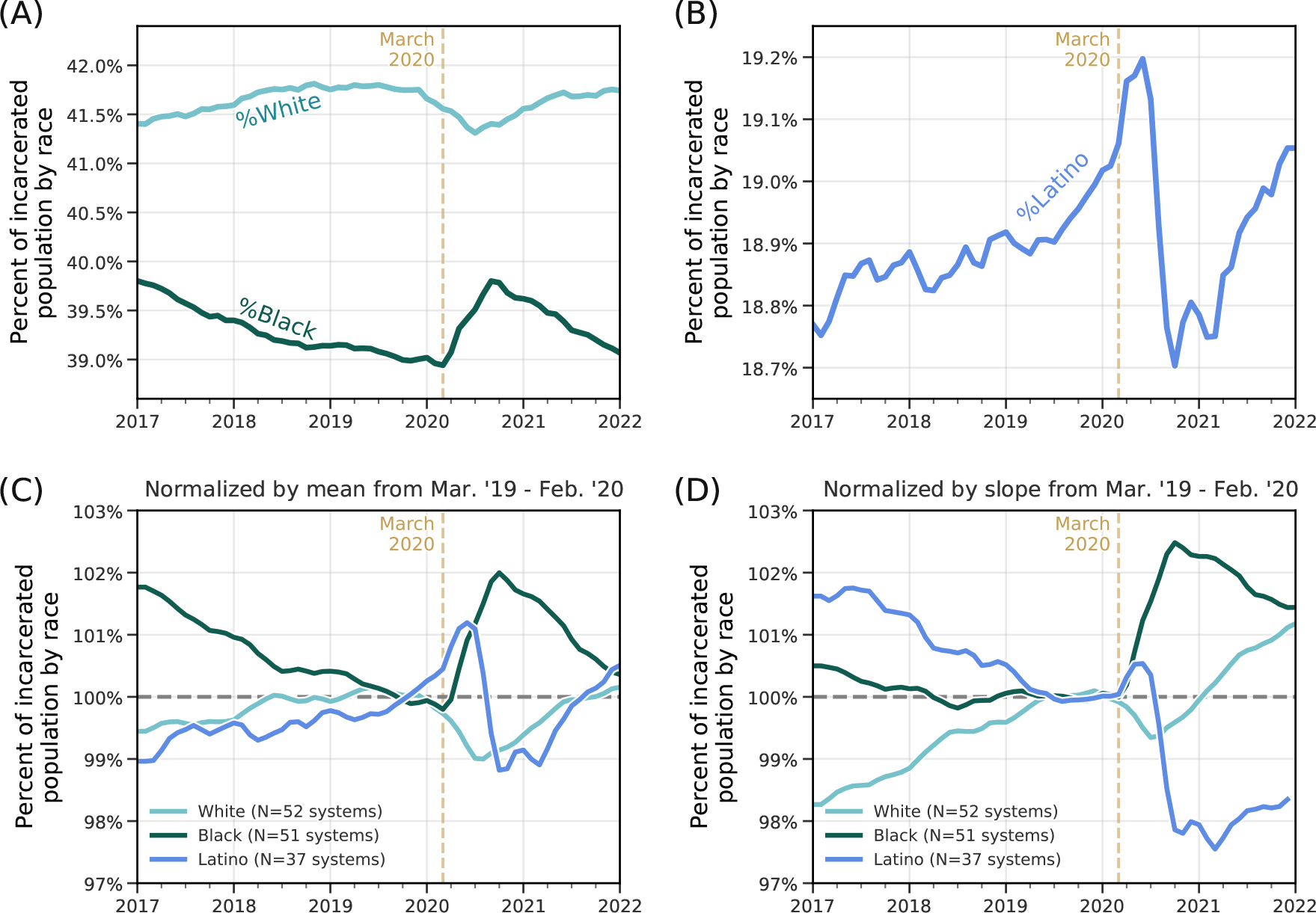
Comparison of Black, White, and Latino incarcerated populations over time. (A) Percent of incarcerated population who are Black and White. Note especially that the effect size of the demographic changes during the COVID-19 pandemic are more pronounced in the incarcerated Black population—see Figure A.16 for additional comparisons. **(B)** Percent of incarcerated Latino people. **(C)** Percentage of White, Black, and Latino incarcerated populations, normalized by the average value between March 2019 and February 2020. **(D)** Percentage of White, Black, and Latino incarcerated populations, normalized by the slope of each curve between March 2019 and February 2020.

**Figure 4:**
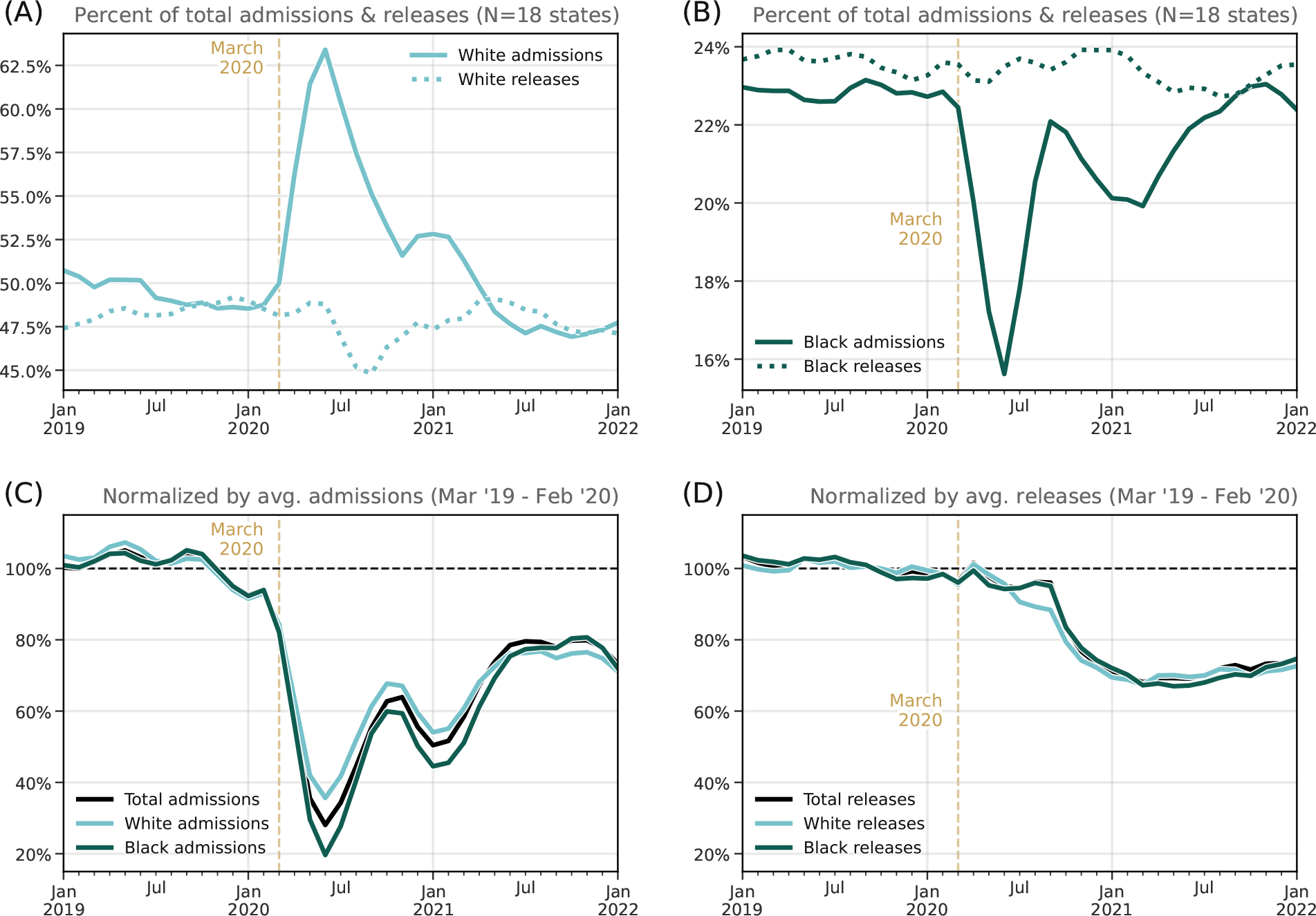
Comparison of admissions and releases by race for 18 states. While data on monthly admissions to and releases from prison are less readily available than prison population data, we can nevertheless highlight the average dynamics of 18 states’ data. **(A)** Percent of total monthly admissions (solid) and releases (dotted) who are white. **(B)** Percent of total monthly admissions (solid) and releases (dotted) who are Black. **(C)** Normalized comparison of the change in monthly white/Black/total admissions. **(D)** Normalized comparison of the change in monthly white/Black/total releases.

The policies, societal disruptions, and behavioral changes that emerged following the onset of the COVID-19 pandemic amplified existing and long-standing racial disparities in the U.S. carceral system. Consistent with other research, we find that disparities in sentencing by race are core to maintaining structural inequalities in incarcerated populations [19, 28].

## 3 Discussion

After declining steadily for the last decade, the percent of Black and other non-white incarcerated people increased sharply during 2020, a trend that was present in almost every prison system across the country. To identify the mechanisms behind this increasing racial disparity, we collected and validated an unprecedented dataset that includes state-level information on police encounters, court proceedings, and incarcerated populations. In order to obtain such granular information across all 50 states, the District of Columbia, and the Federal Bureau of Prisons, we manually collected data from individual Departments of Corrections and filed numerous Freedom of Information Act requests (see Data & Methods).

Prior to the COVID-19 pandemic, racial disparities in admissions were smaller than disparities in the prison population, as recent trends show a migration towards a class-driven disparity, with lower-educated white people steadily increasing in their rate of admission [29]. In a sense, courts had been serving as an instrument for decreasing the racial disparity in prisons prior to the pandemic; for example, the Black-white disparity in prison admissions is typically a ratio of 2:1, whereas it is closer to 6:1 for the total incarcerated population [19] (see [30] for a recent exploration of several factors underlying this trend). Thus, when court proceedings or transfers from county jails are disrupted (i.e., admissions are reduced), the racial disproportionality in the total prison population accelerates, as observed in Figure 1, 2, A.1, and A.2. These dynamics, happening within prison systems nationwide that sees incarcerated Black and other non-white people sentenced for longer periods of time on average [28], led to the abrupt nationwide increase in the percent of incarcerated Black people, starting in March 2020. Differences in the length of sentence by race appear to be a key factor in producing the trends from Figure 1B, but this effect will then be compounded if—in addition to overall decreases in admissions—there are also sudden changes in the typical distribution of the race of people admitted into prisons, which we see, for example, in Texas during the summer of 2020 (Figure A.20B).

Understanding the role that racial disparities in sentencing play in producing the trends from Figure 1B is key for making predictions about how sudden societal disruptions or policy changes in the future may impact prison population demographics (e.g. continued pandemic, Supreme Court decisions, widespread social protests, etc.). These findings can, in turn, help inform policy reform efforts. The sentencing disparity mechanism described in this work is even useful for explaining the dynamics behind the five states that did not conform to the overall national trend in Figure 1B (Maine, Maryland, Missouri, Oregon, and Wyoming; see Figure A.4). These five states maintain prison systems that incarcerate, on average, fewer people under shorter (less than 2-year) sentences, according to data from the National Corrections Reporting Program [31] (differences explored in Figures A.5 and A.6). This observation that states with fewer short term prison sentences did not show the same racial disparity we found nationally, has two subtle but important consequences: First, it suggests that a key reason why the disparities emerge is due to releases of incarcerated people who served shorter-term sentences (without a corresponding amount of admissions). This makes sense, because on any given day, a randomly-selected person being released from prison is likely to have been sentenced for a shorter time period. Second, if white people are more likely to serve shorter sentences, then an overall reduction in the amount of people serving shorter-term prison sentences means there are fewer people serving shorter-term sentences who could be “eligible” to drive the main effect in Figure 1.

While racial disparities in sentence lengths appears to be the most robust explanation behind the trend in Figure 1B, we want to avoid disregarding the potential effects that racial disparities in prison admissions could have played during the COVID-19 pandemic. For example, there is another well-known mechanism through which court closures could have affected different states’ relative rates of Black and Latino prison admissions during the pandemic (see Figure A.18): relative increases in pre-trial case dismissals (Figure A.18D) and pre-trial plea deals. Plea deals in particular have long been demonstrated to result in a disproportionate number of Black defendants spending time in prison [12, 32, 33]. Interruptions in court proceedings may have contributed to the increased Black and Latino representation in prisons populations by 1) reducing the increasingly large flux of white prison admissions, 2) amplifying processes—pre-trial case dismissals and pre-trial plea deals—that are long understood to be a leading contributor to disparities in judicial outcomes for Black individuals. Disruptions in the typical, pre-pandemic court proceedings also offer a compelling explanation as to why (as seen in Figure 1B) we see the reversion to pre-2020 levels, starting in early 2021: the reduction in admissions stopped and, in most states, the total incarcerated population began to increase once again (see Figures 2 and A.1).

Beyond disparities in sentencing and admissions, the COVID-19 pandemic provided several specific challenges that shaped release patterns. Maintaining the largest and most expansive prison system in the world is a major challenge to public health, especially in the context of infectious diseases [4, 34]. In particular, severely overcrowded conditions have presented a public health threat during the COVID-19 pandemic [35]. The physical and administrative structure of prisons provided constraints on ways to quarantine incarcerated people and de-densify congregate settings [35–39]. In recognition of these circumstances, several states enacted policies and initiated executive orders to release individuals who they deem eligible [23]. As a public health intervention, decarceration is a highly effective way to mitigate outbreaks inside and outside of prisons [4, 35, 37–42]. During the pandemic, criteria for decarceration differed from state-to-state, but often included factors such as the age of the incarcerated person and the offense for which they were convicted (e.g. nonviolent drug offenders) [43]. We were able to quantify disparities in the some states’ efforts to de-densify prisons (e.g. in Arkansas, see Section A.4.3 and Figure A.23), which suggests that even decarceration policies widely understood to be consistent with effective and ethical public health practice (and that are assumed to be “race blind”) are susceptible to existing structural and racial inequalities. And one of the most important consequences of disparities in releases is not only about who is released, but who is left behind: the increase in the proportion of incarcerated Black and other non-white people translates to their being at a heightened risk of exposure to SARS-CoV-2.

Taken together, our findings reveal that the pandemic provided a “stress test” for the criminal legal system. In engineering, stress tests involve exposing an apparatus to extreme conditions in order to reveal its fragilities; under these conditions, it can be easier to uncover the underlying mechanisms that govern it. Using a range of data sources, we have argued that COVID-19 amplified underlying racial disparities in the carceral state. As is the case with many complex systems, the dynamics of prison populations are defined by interactions between multiple actors that, in combination, create surprising or troubling results. In response to these findings, society has an ethical obligation to act, and reform sentencing practices and the broader criminal legal system towards more equitable ends.

## Data Availability

The incarceration data used in this work are public records in each state, and we have included the source urls in Table A.1. Together, data from all 50 states, the District of Columbia, and the Federal Bureau of Prisons create The Dataset on Incarcerated Populations, which we have made publicly available via an archived Zenodo repository [56] and a Github repository (https://github.com/jkbren/incarcerated-populations-data). The source data used to construct the Dataset on Incarcerated Populations is available via direct download through the links provided in Table A.1, by public records request, or by request to the corresponding author(s).

## 4 Data & Methods

### 4.1 State prison populations over time

Time series data about states’ prison population over time were collected manually through scraping Departments of Corrections websites, as well as direct requests to state officials through public record requests (e.g. Freedom of Information Act requests, etc.). For every state in our dataset we sought the most temporally resolved data as possible. We collected population data at either weekly, monthly, quarterly, or, for some states, yearly levels. The most common form of data we were able to collect is the number of currently incarcerated persons in a given state, on a monthly timescale. In Table A.1, we link to the data source for every state in our dataset, and in Section A.1, we show how the prison population of every state has changed over time.

We compared the data collected here to data from other organizations that report statistics about the U.S. prison population—the Bureau of Justice Statistics (BJS) and the Vera Institute for Justice [44]—and find high overlap between all three of the datasets. In Section A.3, we identify where our data differs from the BJS data, and we offer an explanation for why we are confident in accuracy of our approach (e.g. in several cases, we received the data directly from the states’ Departments of Corrections, via public records requests).

For every state in this dataset, the total prison population includes both male and female incarcerated people (something that is not always the case in studies about the U.S. carceral system, which so often focuses on male incarcerated people). In New Mexico, Vermont, and California, “Transgender”, “Other”, or “Non-Binary” are also listed as gender categories, though this practice is not widely adopted in reporting statistics about the incarcerated population. In 27 states, incarcerated race statistics are separated by “male”, “female”, and “total”, and further characterizing the interaction between race and sex in biases in admissions and releases during the COVID-19 pandemic remains future work.

### 4.2 State policy data

#### 4.2.1 Court closures and reduced admissions

Qualitative data on the closure and reopening of all 50 state court systems were collected primarily through the administrative orders and/or press releases of each state system’s Supreme or Superior Court or chief judicial officer as well as through local news coverage. The vast majority of states suspended all in-person proceedings with the exception of limited emergency matters between March 12 and March 20, 2020. Several states that adopted policies early in this period issued increasingly strict guidance as the pandemic worsened. New Jersey, for example, suspended new trials on March 12 and issued a two-week suspension on municipal court proceedings on March 14 before finally suspending all proceedings (with emergency exceptions) on March 15. In addition to closing judicial buildings and suspending proceedings, most court closures also extended statute of limitations and filing deadlines due to pandemic disruption. A handful of states, Pennsylvania and Texas among them, permitted or encouraged courts to begin conducting remote proceedings in their initial closure orders, though the adoption of remote proceedings was not widespread in this initial lockdown stage. Court reopening policies were significantly more heterogeneous than the initial closures, though trials remained suspended in most states through at least early-Summer 2020 (and in most cases substantially later). The earliest such policies appeared at the beginning of April 2020, with most aimed at giving regional and local judges discretion to begin hearing proceedings remotely (e.g. Louisiana, Massachusetts, Florida, Iowa, among others). A substantially larger group of states adopted reopening guidelines between late-April and mid-May, many of which allowed essential judicial staff to return to offices following new public health guidance while also maintaining remote proceedings and expanding the number of non-trial proceedings that courts could conduct remotely. Further reopenings and the resumption of limited in-person proceedings took place in many states throughout June, July, and August 2020, though trial proceedings remained suspended. Notably, several states, especially those that adopted phased reopening plans, restricted in-person proceedings and further delayed trial resumption with the Fall-Winter 2020-21 COVID surge. In many states, most administrative orders restricting court operations have at the time of publishing been rescinded, though others, California notably among them, still retain certain accommodations including the option for remote proceedings.

#### 4.2.2 Release policy data

Data on COVID release policies, where they existed, were collected from states’ individual corrections/prison bureau systems, governors’ executive orders, and local news coverage. Fifteen states did not adopt any official release policy, though our data nevertheless shows that there were still reductions in the overall prison population during the pandemic in all of these states. The remaining 35 states adopted policies with varying degrees of specificity and effectiveness, though many overlapped in their broadest contours, allowing consideration for early release to be granted to incarcerated people at increased public health risk (either due to age or underlying health condition) and for those nearing parole and/or the end of their prison sentences.

Almost all states with such policies did, however, adopt a restriction preventing the release of those incarcerated for violent crimes or sex offenses. North Dakota was an outlier in this regard. Of the 120 people the state initially released from prison in March 2020, 14 were serving time for violent crime convictions and 11 were convicted of sex offenses. New York’s release policy was notably more restrictive (on paper at least) than many other states—only those incarcerated for “non-criminal technical parole violations” were eligible for COVID release. As an example of one state’s release policy, we include below an excerpt from the Virginia Department of Corrections’ policy on releases [45], from April 24, 2020:

The Director of the Department of Corrections is authorized to consider early release for individuals with less than one year left to serve while the COVID-19 emergency declaration is in effect. Offenders convicted of a Class 1 felony or a sexually violent offense are not eligible for consideration. The exact number of individuals eligible for early release consideration will change depending on the length of the emergency declaration order. The [Department of Corrections] will identify those that are eligible for consideration using the procedures it has developed to ensure public safety and will notify offenders who are to be released under the early release plan. A diagnosis of COVID-19 is not a release factor.

The following Early Release Criteria will be utilized in considering an incarcerated person for early release pursuant to legislation:

- **Release Date**: The inmate’s Good Time Release Date must be calculated and verified in order for the incarcerated to be considered.
- **Inmate Medical Condition**: The inmate’s medical condition will be considered.
- **Offense History**: By legislative mandate, early release does not apply to inmates convicted of a Class 1 felony or a sexually violent offense. Consideration for early release will be based on the seriousness of the current offense, in descending order as follows: Non-violent Offense, Felony Weapons Offenses, Involuntary Manslaughter, Voluntary Manslaughter, Robbery, Felony Assault, Abduction, Murder, Sex Offense
- **Viable Home Plan**: The incarcerated person must have a documented approved home plan to be considered.
- **Good Time Earning Level**: The inmate’s current good time earning level must be I or II to be considered.
- **No Active Detainers**: Inmates must have no active detainer to be considered.
- **No Sexually Violent Predator Predicate Offenses**: Inmates convicted of one or more sexually violent offenses established in §37.2-903 of the Code of Virginia are not eligible pursuant to legislation.
- **Recidivism Risk**: Inmates must have a risk of recidivism of medium (5-7) or low (1-4), as identified by the validated COMPAS instrument, to be considered.

Note especially the inclusion of the COMPAS risk assessment tool, which is used in court systems across the U.S. as a way of quantifying an offender’s likelihood of re-offending (recidivism). Over the last several years, we’ve seen a growing body of scholarly work devoted to identifying problematic and harmful racial and economic biases that arise when algorithmic risk assessment tools are used in practice [46–51]. COMPAS, in particular, has been the subject of a number of studies that take a critical look at the effectiveness—and ethics—of these risk assessment tools in the justice system [47, 52]; in one study, COMPAS was found to predict recidivism 61% of the time, but at the same time, Black people were almost twice as likely to be labeled as high risk for re-offending but not actually re-offend [52].

Further research is needed to quantify demographic patterns in the incarcerated individuals who were released across different states, and because there was such high heterogeneity in different states’ policies, it remains an open question whether we will see the same broad, systematic racial differences among the people who were released. However, as has been the case throughout the COVID-19 pandemic, heterogeneous policy responses across localities has typically had detrimental effects on our collective response to the pandemic [53].

### 4.3 Study definitions of race and ethnicity

The data that we collected for the study used definitions of racial and ethnic groups that were determined by the agencies that collected the data. When the authors are discussing race and ethnicity in their interpretations, they are referring to the historical categories that have social, cultural and political consequences. We use the term “Latino” to describe persons that are otherwise described as “Hispanic” in many settings. We have used the term “non-white” in select locations, as not all states had data disaggregated into the same set of categories. And so for some analyses, “non-white” directly describes the available data. For a table of the race categories reported by every state in our dataset, see Table A.7.

Recent advances in medical conventions have prompted discipline-wide introspection about the ways that race and ethnicity are discussed and used in research [54]. This is of critical importance to health equity and racial justice, and while in this work we rely on race statistics reported by states’ Departments of Correction, future work will critically examine the differences in approaches for reporting race and ethnicity statistics of incarcerated populations. Notably, it is important to know whether a state’s statistical reports use race categories that have been self-reported by the incarcerated person or whether it is interviewer-observed, which is often the case in administrative databases. These approaches are quite different and often result in inaccuracies in measurement of racial disparities [55]. Lastly, in Section A.3.3, we introduce a novel dataset that contains policies from 48 states and the Federal Bureau of Prisons about whether race data of incarcerated individuals is obtained via self-report or visual-assignment from administrators or staff.

## Additional information

### Acknowledgements

The authors thank Harrison Hartle, Stefan McCabe, Timothy LaRock, Ryan Gallagher, Morgan Frank, Rori Rohlfs, Elizabeth Ross, Brandon Terry, Sharad Goel, Danielle Allen, and members of the Northeastern University NULab for helpful conversations and tips with constructing the dataset. The authors would like to thank members of the Justice Collaboratory at the Yale Law School, the Center for the Study of Policing, Incarceration, and Public Safety at The Hutchins Center for African & African American Research, Harvard University, the MLK Visiting Scholars Program at the Massachusetts Institute of Technology, and the Vera Institute for Justice for support, helpful exchanges on the subject matter and feedback on the manuscript. Lastly, the authors would like to thank the Santa Fe Institute, New York University, University of Pittsburgh, and the Yale School of Public Health for seminar and colloquium invitations, where aspects of this study were discussed.

### Code availability

The Python code to reproduce the analyses and construction of the database is available at https://github.com/jkbren/incarcerated-populations-data and [56]; this repository contains several Jupyter notebooks with analyses and tutorials on how to automate the collection of some of the data used here.

### Data availability

The incarceration data used in this work are public records in each state, and we have included the source urls in Table A.1. Together, data from all 50 states, the District of Columbia, and the Federal Bureau of Prisons create the “The Dataset on Incarcerated Populations”, which we have made publicly available via an archived Zenodo repository [56] and a Github repository (https://github.com/jkbren/incarcerated-populations-data). The source data used to construct the Dataset on Incarcerated Populations is available via direct download through the links provided in Table A.1, by public records request, or by request to the corresponding author(s).

### Competing interests

The authors declare no competing interests.

### Author contributions

B.K., C.B.O., S.V.S., & E.H. conceived the project. C.B.O., S.V.S., & E.H. directed the project. B.K. directed the construction of the data science pipeline. B.K., B.J.S., Z.B., P.K., J.S., A.S., & N.K. collected data. B.K., C.B.O., B.J.S., and S.V.S. conducted analyses. B.K., C.B.O., E.A.W., T.E.R., S.V.S., & E.H. interpreted and integrated the results. B.K., C.B.O., B.J.S., E.A.W., T.E.R., S.V.S., & E.H. contributed to researching, writing, and editing the final manuscript.

## A Supplemental Information

### A.1 State-by-state breakdowns

In Figure A.1, we show the prison population over time for all 50 states, the District of Columbia, and the Federal Bureau of Prisons. In Table A.1, we give an overview of the scope of each state’s data in our dataset. In Table A.2, we list every state in order of the maximum reduction in prison population, alongside the month that this decrease was observed. In Figure A.2, we reproduce state-specific versions of Figure 1B.

Note that for several states in Figure A.2, we plot the percent incarcerated population who are not white, as opposed to Black. This is due to either the small number of incarcerated Black people in the state (as in Hawaii, Idaho, South Dakota, Vermont, etc.) or the absence of Black as a racial category in the state’s reports (e.g. Michigan). With this state-by-state view, we see that the only state that unequivocally does not show a similar trend to Figure 1 (and in fact, shows the opposite) is Oregon; ongoing and future work will attempt to disentangle whether this is a reflection of the state’s criminal justice policies, demographic patterns, reporting procedures, or any of the above.

### A.2 A nationwide trend or state-level heterogeneity?

In this section, we report raw and interpolated data for each state, the District of Columbia, and the Federal Bureau of Prisons, in order to compare time series of the percent Black population in prisons to the percent non-white population (see [4]). In some cases, the main effect observed in Figure 1B is clearly recapitulated; in others, we do not see the effect in the percent Black population, but rather we see it in the percent non-white population. The only states where we do not observe the main effect in both demographic categories are Maine, Oregon, and Wyoming.

In Figure A.3, we plot the distribution of the slopes of each state’s percent incarcerated Black population data during different time windows (2017-2019, 2019-2020, 2020-2021, and 2021-2022). With few exceptions, this plot shows that the slope of the percent Black incarcerated curves increased during 2020, compared to the 2019 averages. After this, the same slopes decreased during 2021. Isolating these slopes is useful again for nationwide comparison for two reasons: 1) because it can allow us to ask questions about what we would expect to happen if we did not observe some sort of spike in the percent Black population in 2020-2021, and 2) because we can use the slope of 2019-2020 as a way of normalizing the entire time series, allowing for a simple test of whether or not we observe a spike in 2020.

Figure A.4 normalizes each state’s percent incarcerated Black population data by the best-fit line from its *slope* between 2019-2020. That is, we divide each state’s time series of percent Black population by the corresponding value in the best-fit line from the slope from the year prior to March 2020. Visually, this bundles the states’ curves to the 100% value before the pandemic (i.e., 100% of normalized pre-pandemic values). After March 2020, there are five states that do not at least briefly show a spike in their percent Black population: Maine, Maryland, Missouri, Oregon, and Wyoming.

#### A.2.1 Interpolating monthly population data

In [4], we have included tables of the population and demographic data for each prison system studied here. Also included in these tables is a tag about whether or not the data is raw data from the state or interpolated. For most (35) prison systems, we have raw monthly data for the entire duration of the study period. For some, we only have data at the quarterly (8) or bi-annually (4); for these 12 states, we simply do a linear interpolation on the raw demographic data and sum these columns together to arrive at a total estimate for the number of people incarcerated each month in between the quarterly/biannual dates. As a validation, we do this same interpolation on states where we do have reliable monthly data, and the estimates are almost perfectly aligned (average *R*^2^ of correlation with ground truth for bi-annual interpolation: 0.975; annual interpolation: 0.945). We show this high correspondence for three example states in Figures A.11, A.12, and A.13. For five states: Michigan, New Jersey, South Carolina, Tennessee, and Virginia, we only have demographic data at the yearly level. In each of these states, we have population totals at the monthly level; with these, the task becomes to estimate the counts of incarcerated people by race each month, given the population totals. In some ways, this is an easier task, since we know the overall trend in the prison population. Here, again, we do a linear interpolation between the dates without missing values, multiplied by a factor of (interpolated_sum / actual_sum). Doing this same validation on states with reliable, monthly data reporting gives us high alignment again. Lastly, we note that every combination of including or excluding states based on their reporting frequency and quality still produce the same qualitative results, which we would expect given the extensive discussion above.

### A.3 Comparison across prison population datasets

Other organizations collect and report data about prison populations over time. In order to situate the data used here within a broader body of work studying U.S. prison population trends, we validate against data released by the Bureau of Justice Statistics [2] (BJS) and the Vera Institute for Justice [3] (Vera). In Figure A.10, we plot the BJS’s yearly estimates of the number of people in state prisons across the United States from 2014 until 2020. We concatenate the BJS data with the Vera data to approximate a “ground truth” estimate for the prison population over time.

We note several key points. First, starting in 2020, our dataset almost exactly matches the Vera dataset. Prior to 2020, our dataset reports a prison population that is approximately 1% smaller than the BJS data. After investigating what could have brought about these differences between the two datasets, we identified five states with the largest between-dataset differences (Montana, Florida, Texas, Virginia, and Ohio; see Table A.3). Because of these discrepancies, we took additional care to confirm that the data we had collected was exactly what was reported by states’ Departments of Corrections websites (or sometimes, through Freedom of Information Act requests). In Section A.3.1, we describe the rationale for why we are confident in the data included in the present study, and we also directly link to the data sources used to offer transparency in the data collection process.

#### A.3.1 Comparison to Bureau of Justice Statistics data

To our knowledge, the scale of the data that we assembled in this work is unique among the available public datasets about states’ prison populations over time. In Table A.3, we dive deeper into the discrepancies between the data used here and those that were released by the Bureau of Justice Statistics [2]. We offer explanations that reconcile why we may observe such differences, and we conclude that the data reported here is consistent with state prison population statistics reported by states’ Department of Corrections.

#### A.3.2 Comparison with the National Corrections Reporting Program

We drew on another large, well-known dataset to validate the findings from the data we collected: the National Corrections Reporting Program (NCRP) [1]. The NCRP data we used contains detailed information about individuals incarcerated in almost every state, yearly, until December 31, 2019. In recent years, data from the NCRP has been subject to scrutiny both in terms of its coverage and completeness as well as how it reports data about the race of incarcerated people [5–9].

Nevertheless, these data are a cornerstone of legal and justice research in the United States, and as such, we sought to use it as a benchmark of pre-2020 data to see 1) the extent to which there is significant overlap between the two datasets and 2) if we analyzed the subset of our data that only had high overlap with the NCRP data (i.e., exclude states where there is significant disagreement between the two datasets), whether we would reproduce the main result in Figure 1B.

In Figure A.14, we plot a comparison between the NCRP data and our data for 12 states’ total incarcerated population over time (NCRP data plotted in red). Immediately, we are reassured about the correspondence between these states in the two datasets. There are, however, several states where the overall trend in the prison population is similar between the two datasets, just shifted uniformly up or down (i.e. states with the same or similar-shaped curves but shifted by a fixed amount). Lastly, there are several states where the NCRP data is clearly not capturing the same information that our dataset contains. These are states that—we suggest—do not have high coverage or high data quality in the NCRP dataset or have otherwise changed their reporting protocol during the duration of their inclusion in the NCRP.

Ultimately, if we only analyze states with high overlap between NCRP and our data (a proxy for overall reporting quality: Washington, California, Nevada, Utah, Nebraska, Arizona, Colorado, Wyoming, Kansas, Texas, Iowa, Minnesota, Illinois, Indiana, Kentucky, Tennessee, Mississippi, West Virginia, Ohio, Wisconsin, Georgia, Florida, New Jersey, New York, and South Carolina), we see the same qualitative result (see Figure A.15). We see this as a validation with multiple benefits: First, it grounds the data we have collected in a known to a well-studied companion dataset. Second, we see an opportunity to use our data to augment or help fill in states with known reporting irregularities or other issues in the NCRP dataset. While the insights from these comparisons between the two datasets were not the intended contribution of our paper, we are encouraged nonetheless that these analyses offer a roadmap for future work improving data quality in the NCRP.

#### A.3.3 Survey of states’ race reporting procedure

As briefly mentioned in the previous section, there remain several known challenges in relying on administrative data to study racial disparities. In particular, one common issue is the heterogeneity between how states and other government agencies report data on race. Broadly, there are two main approaches to collecting information about an individual’s race and/or ethnicity: self-reported from the individual in question or “visual inspection” by either a clerk, law enforcement official, or other administrative staff. On the one hand, some scholars argue that external assignment of race more accurately reflects the scale at which discrimination or prejudice operates [10] and should therefore be relied upon for studying race and ethnicity. On the other hand, self-reported data about an individual’s race is likely more accurate and therefore more useful for large-scale quantification [11, 12]. Many researchers argue for an array of different survey questions in order to accommodate both approaches [13, 14], while others have found ways of improving the accuracy of administrative race data through a variety of post hoc statistical corrections [15, 16]. An Oregon Criminal Justice Commission report, for example, analyzed race/ethnicity data in their management system and discovered that the system regularly mis-labeled individuals’ race, such Latino, American Indian, and Asian individuals were under-represented by up to 15% [17]—a number that can be corrected through statistical re-weighting. Table 5 of the BJS *Prisoners in 2020* report emphasizes this issue and describes the steps taken to statistically adjust the data in order to estimate the underlying racial distribution in state and federal prisons [18]:

“National-level estimates of the number of persons by race and ethnicity under the jurisdiction of state prisons on December 31, 2020 were based on an adjustment of NPS [National Prisoner Statistics] counts to comply with the Office of Management and Budget (OMB) definitions of race and ethnicity… Not all NPS providers’ information systems categorize race and ethnicity in this way. In addition, these data are administrative in nature and may not reflect a prisoner’s self-identification of race and ethnicity… For state prisoners, BJS calculated the ratio of the distribution of state prisoners by race and ethnicity in BJS’s selfreported prisoner surveys, which use OMB categories for race and ethnicity, to the distribution of prisoners by race and ethnicity in NPS data for the year closest to the fielding of the survey. BJS then multiplied this ratio by the distribution of state prisoners’ race and ethnicity using the current year’s NPS. The percentage of persons self-reporting to the NPS as non-Hispanic and as two or more races was assumed to be equal to that of the self-reported prisoner survey. The final percentage distribution of race and ethnicity was multiplied by the total of sentenced state prisoners to obtain counts for each category.”

In other words, the BJS performs a statistical correction that attempts to correct for heterogeneity in how race is reported across states. A natural question that arises here is whether we think that the same heterogeneities in administrative race reporting are present in the dataset we introduce here. If there were systematic differences in how race is reported by state (e.g. some states may report the race of a newly incarcerated person via a self-reporting procedure during intake; other states may record this data via a staff member assigning a race during intake based on visual features), this could potentially be problematic for the main results in this work. That is, we may be systematically mis-estimating the magnitude of the observed trends in Figure 1B. Alternatively, it may be even more problematic if there are non-systematic differences in how race is reported; in this case, it could potentially require a different statistical correction performed for each state, opening the dataset up to a deserved scrutiny. Fortunately, we do not think the dataset used in this work is subject to these concerns—or if it is, the impact is minimal. The reason for this is based on newly collected data from almost every state’s Department of Correction, displayed in Table A.4^1^.

In nearly every state, it is the stated policy to collect self-reported race data during admissions into prison. There are a few states with either ambiguous policy language (e.g. Minnesota, which explicitly writes “Race information may be self-identified or classified by an observer.”) or policy language that is suggestive of self-report but not entirely (e.g. Massachusetts: “It is mostly self-reported, however, if the county sends a face sheet the Booking Officer will use that.”). One state (Texas) explicitly referred to staff members visually assigning someone’s race “…during intake, the [Texas Department of Criminal Justice] will visually determine the race of the individual.” While Texas was the only state that uses this procedure for collecting race statistics, we must stress that it is the largest prison system in the country, and trends in data from Texas strongly influence national averages.

The other states’ policies are based on self-reported data; we include the precise language from the policy in Table A.4. We do not know of other research with this kind of detailed state-by-state data. Note: here, we do not assume perfect compliance with the self-reporting policy (e.g. either on the administrators or the people being admitted into prison), but at this point, we do see this survey of states’ race data collection policies as the most promising validation of the administrative race data used in this work.

### A.3 Case studies: Court system, policing, and inmate release data

In addition to data about states’ prison populations and prison policy, we also used state-specific data about outcomes of court proceedings, crime/offense type and severity, traffic stops, and inmate releases in order to tell a broader story about the structural effects of mass incarceration during the COVID-19 pandemic.

#### A.4.1 Florida trial statistics data

Using Florida as a case study, we show how changes to typical court proceedings can potentially lead to new racial disparities in the prison population [19–22]. In Figure A.18, we plot monthly trial statistics from circuit criminal defendants in Florida. Prior to March 2020, an average of 14,000 defendants were disposed each month (i.e., pass through the court system and have their charges dropped, agree to a guilty plea, go to a jury trial, or go to a non-jury trial; Figure A.18A). Starting in March 2020, the number of defendants decreased sharply, reaching nearly 4,000 in May 2020, resulting in a backlog of cases (Figure A.18B).

Between March 2020 and June 2020, more than 99% of cases did not go to trial (up from an average of approximately 97% prior to 2020). An increase in the proportion of cases that get resolved pre-trial means that a greater percent of all defendants passing through the Florida courts system will either agree to a guilty plea or see their charges dropped entirely. Importantly, both of these can be sources of statistical bias in the resulting prison population.

First, prior research has demonstrated that Black defendants are almost 70% more likely than white defendants to receive a plea deal that involves spending time in prison [23]. Second, we show here that the percent of cases that were dismissed entirely increased from an average of 10% before 2020 up to 15% in June 2020 (Figure A.18C), and this increase in dismissed cases is strongly correlated with the percent of non-white incarcerated individuals (lagged one month to account for time delays in sentencing; Figure A.18D). This correlation did not simply arise after March 2020—we see these same correlations between percent of dismissed cases and percent of non-white incarcerated people prior to the COVID-19 pandemic, suggesting that this relationship is potentially more general and not merely an anomaly due to the pandemic. This case study highlights multiple potential disparities that can stem from disruptions in the court system, and it emphasizes the need for more states to make similar sentencing data available to the public. In this vein, we report preliminary analyses about the race of defendants whose cases were dismissed in Figure A.19, using data compiled by the Florida Office of the State Courts Administrator (collected by court clerks) to show a relative increase in the number of white defendants among those who had cases dismissed in the early months of the pandemic. See Section A.4.1 for further discussion of these data, including limitations and future work. Having access to standardized data about the race of defendants across every state will further allow us to connect case-dismissals to prison demographic distributions.

To collect the court proceedings data used in Section 2.3 [19], we manually downloaded monthly data about the statewide data on the outcomes of Circuit Criminal Defendants between January 2018 and June 2021. We summarize this dataset in Table A.5.

In addition to summarized monthly statistics about the nature and outcomes of circuit criminal court defendants, we also requested *individual* -level data about the defendants that make up this aggregate data. The Office of State Court Administrator (OSCA) in Florida provided us with data from the Criminal Transaction System for 2018, 2019, 2020, and 2021; these data contain a column for defendants’ race, the action taken by the court, and the date each case was decided (among many other variables)^2^. This dataset allows us to run a simple analysis: Among defendants with cases that were dismissed, between 2018-2021, what percent are recorded as white? We plot this in Figure A.19B (A.19A is a reproduction of Figure A.18C, and A.19C shows the two curves atop one another, rescaled using min-max scaling in order to highlight the timing and relative increase that both measures show after March 2020).

#### A.4.2 Texas offense severity data

In recent years, White/Black/Latino people have accounted a for similar percent of the total incarcerated population in Texas state prisons (33.7%, 32.6%, 33.1%, respectively); mirroring the nationwide trend in Figure 1B, the percent of incarcerated Black people increased sharply in Texas after March 2020 (Figure A.20A). While it is difficult to point to any single cause behind this abrupt disparity, some have argued that more careful consideration must be given to racial differences in the severity of the crimes for which people are incarcerated.

That is, without more extensive data about the incarcerated population in Texas, we cannot rule out the possibility that the observed spike in the relative number of incarcerated Black people is due to a relative increase in the severity of crimes committed, by race. We show, however, that this is not the case.

To do this, we merge data from two sources. The first is monthly data about every incarcerated person in Texas from July 2019 until November 2021 (the Texas Department of Criminal Justice “High Value Dataset” series, from https://www.tdcj.texas.gov/kss_ inside.html); each row in this dataset corresponds to an incarcerated person and includes details about the individual’s race and sex, as well as sentencing information. The second dataset is a table that maps every offense to one of four severity levels: low, moderate, high, or highest (https://www.tdcj.texas.gov/bpp/parole_guidelines/Offense_Severity_ Class.pdf).

We assign an offense-severity category to each incarcerated person by merging the two datasets along the National Crime Information Center (NCIC) code for the sentenced offense. In Figure A.21, we compare these offense-severity categories by race. In Figure A.21A-D, we plot the race distribution within each offense-severity category (i.e., we plot the percent of incarcerated persons with a given offense-severity category who are White/Black/Latino). To accentuate the relative changes in these trends, in Figure A.21E-H, we plot the same curves standardized (i.e., divided by) by their pre-pandemic averages; in these subplots, 100% would indicate no difference from before the pandemic. After March 2020, we see abrupt increases in the relative number of incarcerated Black people in the “low” and “moderate” offense-severity groups (dark green curve, Figure A.21E-F). This is exactly counter to the suggestion that the nationwide trends in Figure 1B are due to Black people committing more severe crimes during the early months of the pandemic. Without these same datasets for every state, we cannot yet say that trends seen in Texas are universal across the United States, but following these analyses, we urge every state to make these types of data available.

#### A.4.3 Arkansas eligible release data

In Figure A.23, we compare the demographics of the prison population in Arkansas in May 2020 to the demographics of incarcerated people eligible for early release under Governor Hutchinson’s authorization [24]; despite the fact that 57.2% of the Arkansas prison population was white, over 72% of the incarcerated people eligible for early releases were white—a disparity that we would not expect to see in a prison system absent of release policies that favored incarcerated white people. These outcomes may manifest in multiple ways: sentencing patterns that create longer sentences for incarcerated Black and Latino individuals, different classifications (e.g., violent or nonviolent) and other categorizations that may drive a disparity in those released.

In Figure A.17 we show that while, on average, incarcerated Black people are released at disproportionately low rates (and incarcerated white people are released at disproportionately high rates), the effect of COVID-19 based releases can temporarily bring about release patterns that are less statistically skewed by race. We see this in Massachusetts especially in May and November of 2020, a sign that in at least one state, it is possible to decarcerate in a way that does not exacerbate existing racial inequalities. A key insight into why this might produce more equitable releases-to-incarcerated ratios in general has to deal with differences in the average length of prison sentence, by race (sentence length, here, we consider to be a proxy—albeit an imperfect one, see [25]—for “time served” in prison). That is, if there are systematic differences in the length of prison sentences by race (e.g. if, on average, Black people in prisons are more likely to be sentenced to longer sentences than white people, which we show in Figure A.22 for Texas), a relative increase in the number of monthly releases would decrease the overall average sentence length, provided that the release criteria is based in part on people who are close to the end of their sentence.

While there are not particularly noticeable changes after March 2020 in the trends of both curves in Figure A.17C, here we directly quantify racial disparities among the inmates who were eligible for release [24]. Using data released by the Arkansas Department of Corrections, we joined inmate identification numbers to their listed race and sex according to the Arkansas Department of Corrections Inmate Search tool (https://apps.ark.org/ inmate_info/index.php).

### A.4 Citation diversity statement

Recent work has quantified bias in citation practices across various scientific fields; namely, women and other minority scientists are often cited at a rate that is not proportional to their contributions to the field [26–33]. In this work, we aim to be proactive about the research we reference in a way that corresponds to the diversity of scholarship in this field. To evaluate gender bias in the references used here, we obtained the gender of the first/last authors of the papers cited here through either 1) the gender pronouns used to refer to them in articles or biographies or 2) if none were available, we used a database of common name-gender combinations across a variety of languages and ethnicities. By this measure (excluding citations to datasets/organizations, citations included in this section, and self-citations to the first/last authors of this manuscript), our references contain 28% woman(first)-woman(last), 22% woman-man, 12% man-woman, 12% man-man, 0% nonbinary, 12% man solo-author, and 14% woman solo-author. This method is limited in that an author’s pronouns may not be consistent across time or environment, and no database of common name-gender pairings is complete or fully accurate.

---

**Figure A.1.**
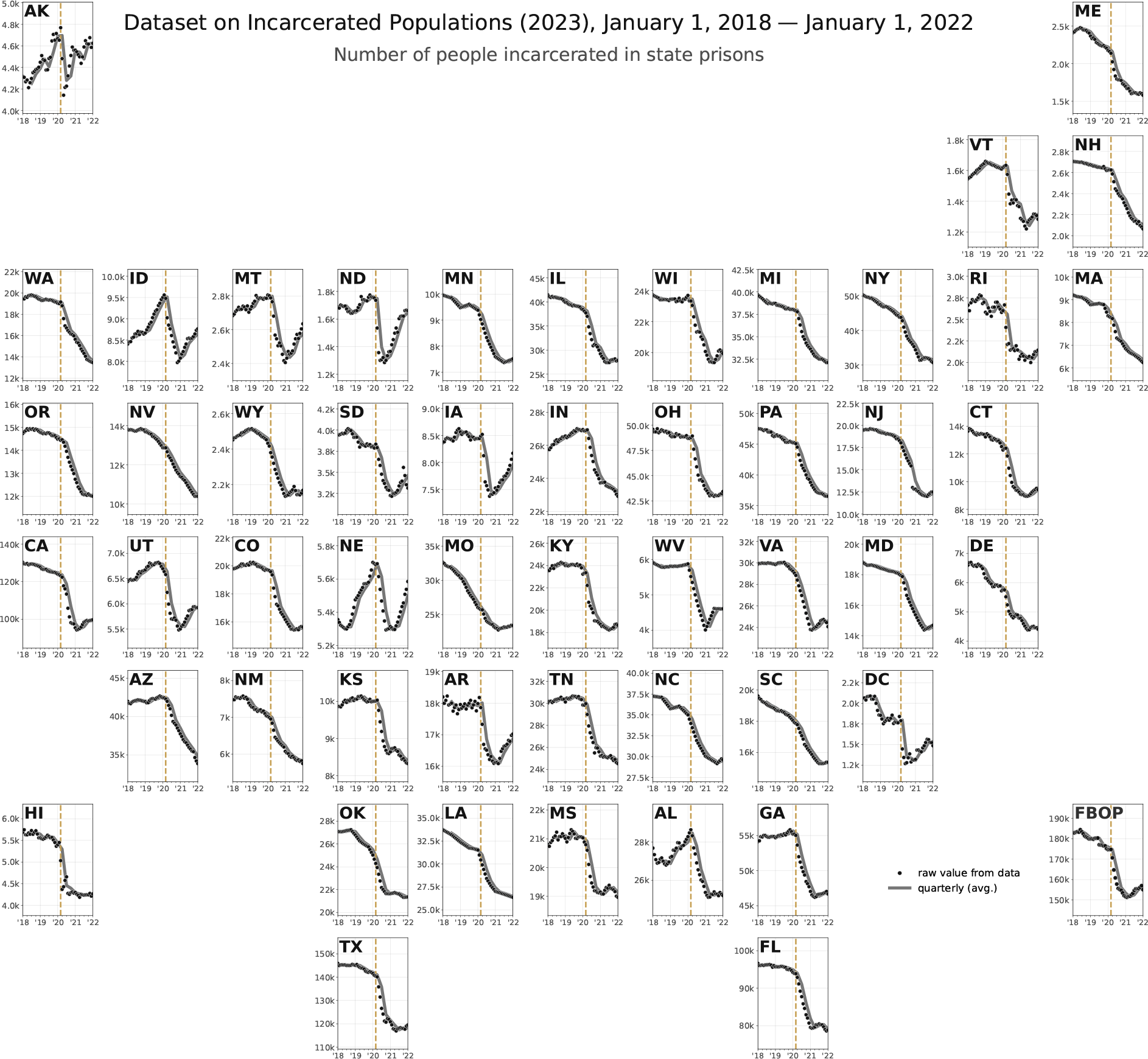
State-by-state (and federal) time series of prison populations. For consistency, the data plotted have been up- or down-sampled to the quarterly level (most states in the dataset report monthly data, but several report twice-yearly and yearly).

**Figure A.2:**
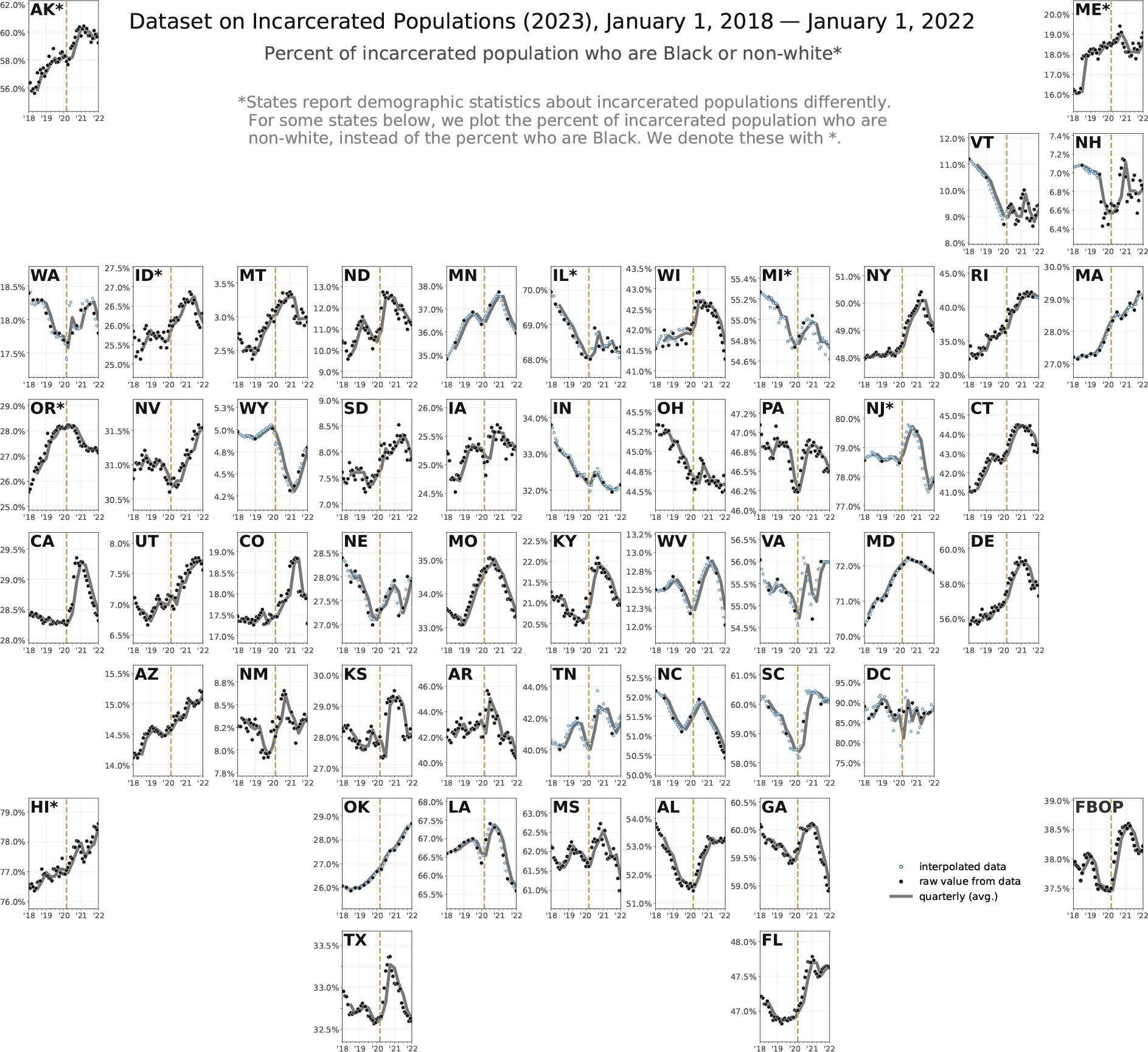
State-by-state (and federal) time series of the percent incarcerated Black people over time. Note: Because of the heterogeneity in reported racial categories and some states’ small incarcerated Black population, we plot the percent incarcerated population who are not white for several states. Vertical axes differ in magnitude by state. For a visualization of the states with the most standardized reporting procedures.

**Figure A.3:**
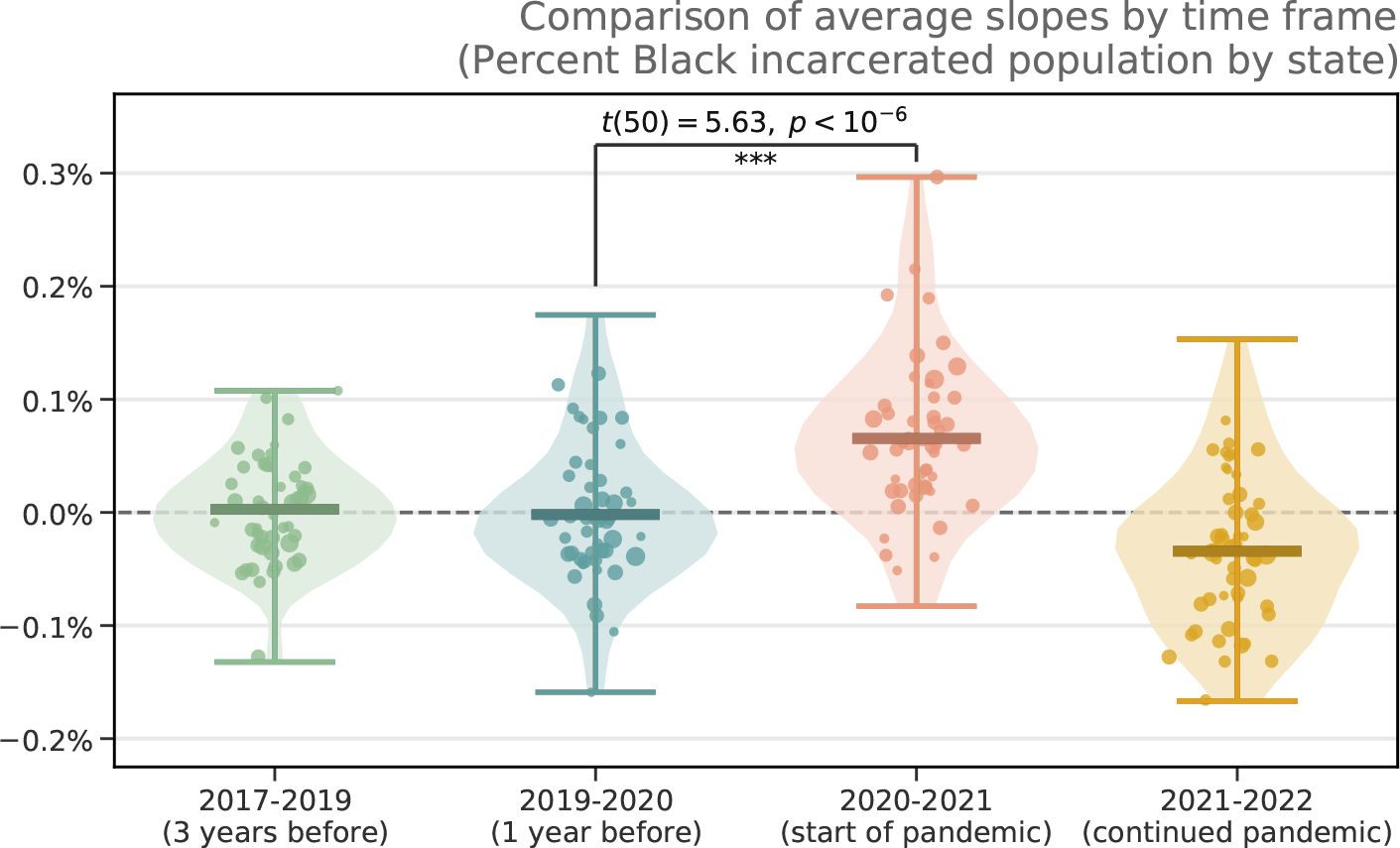
Comparing slopes of percent incarcerated Black population across states. Each violin plot represents every state and federal prison system’s slope of the percent incarcerated Black population curves. Violin plots depict distributions of data, such that the width of each violin corresponds to the density of points in a given range, and the lines at the top/bottom are the max/min of the underlying data. The horizontal lines in the middle of each violin plot corresponds to the means of each distribution. From 2020 to 2021, we observe a statistically significant difference in the slope from before the pandemic (two sided t-test, *t*(50) = 5.6251; *p* = 1.691162 × 10*^−^*^7^).

**Figure A.4:**
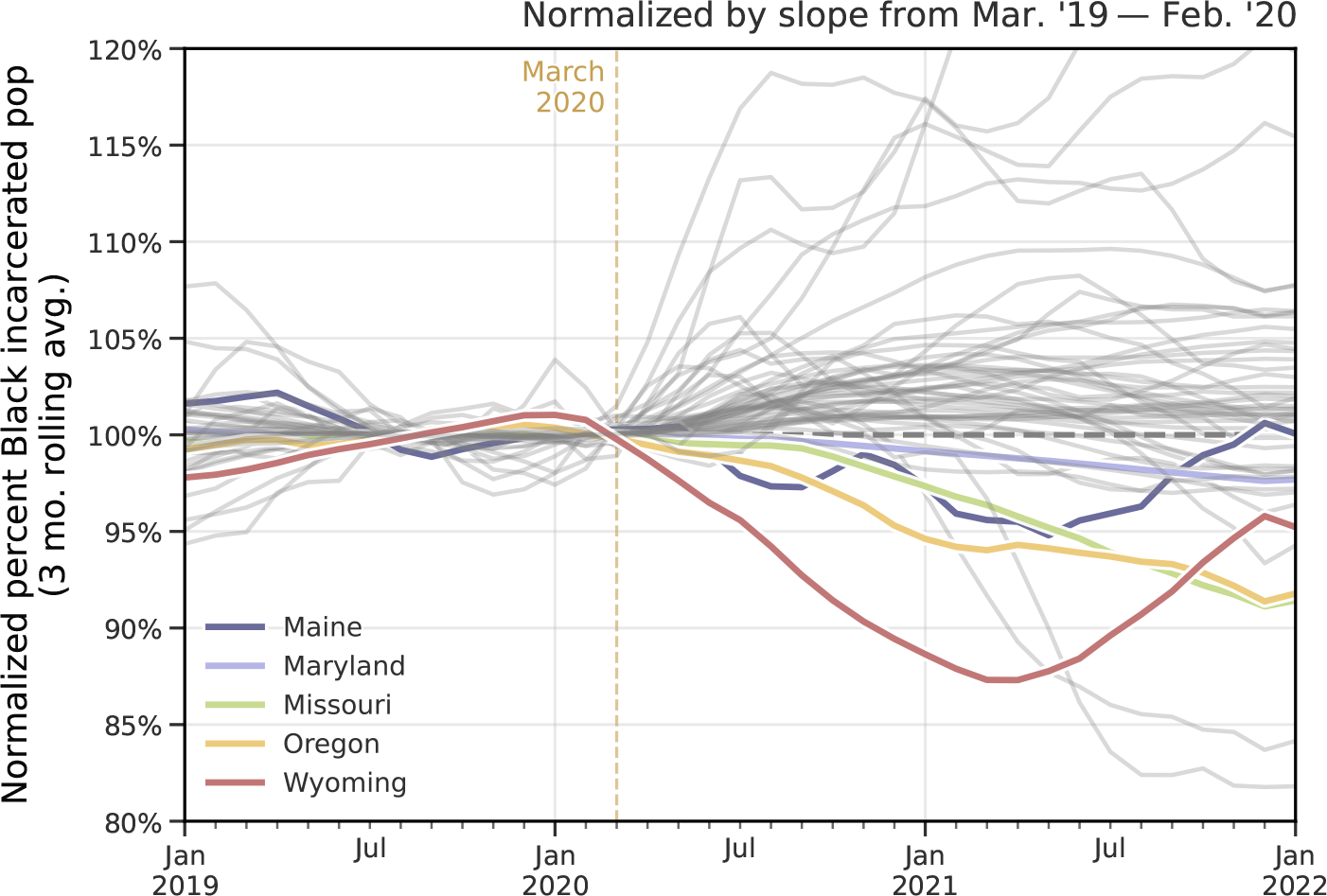
Normalized percent of incarcerated Black population for every state and federal prison system. Applying the same normalization procedure as in Figure 3D (by slope of pre-pandemic curve), we compare the trends of the every state and federal prison system’s percent of incarcerated Black population. Under this view, five states do not follow the broad nationwide trend as in Figure 1B: Maine, Maryland, Missouri, Oregon, and Wyoming. For every other state, we see the percent of incarcerated Black people increase for multiple months, starting after March 2020. The top four curves on the plot correspond to prison systems that are smaller in size (New Hampshire, North Dakota, Vermont, the District of Columbia), with demographic statistics that can be disrupted by small fluctuations in incarcerated populations.

**Figure A.5:**
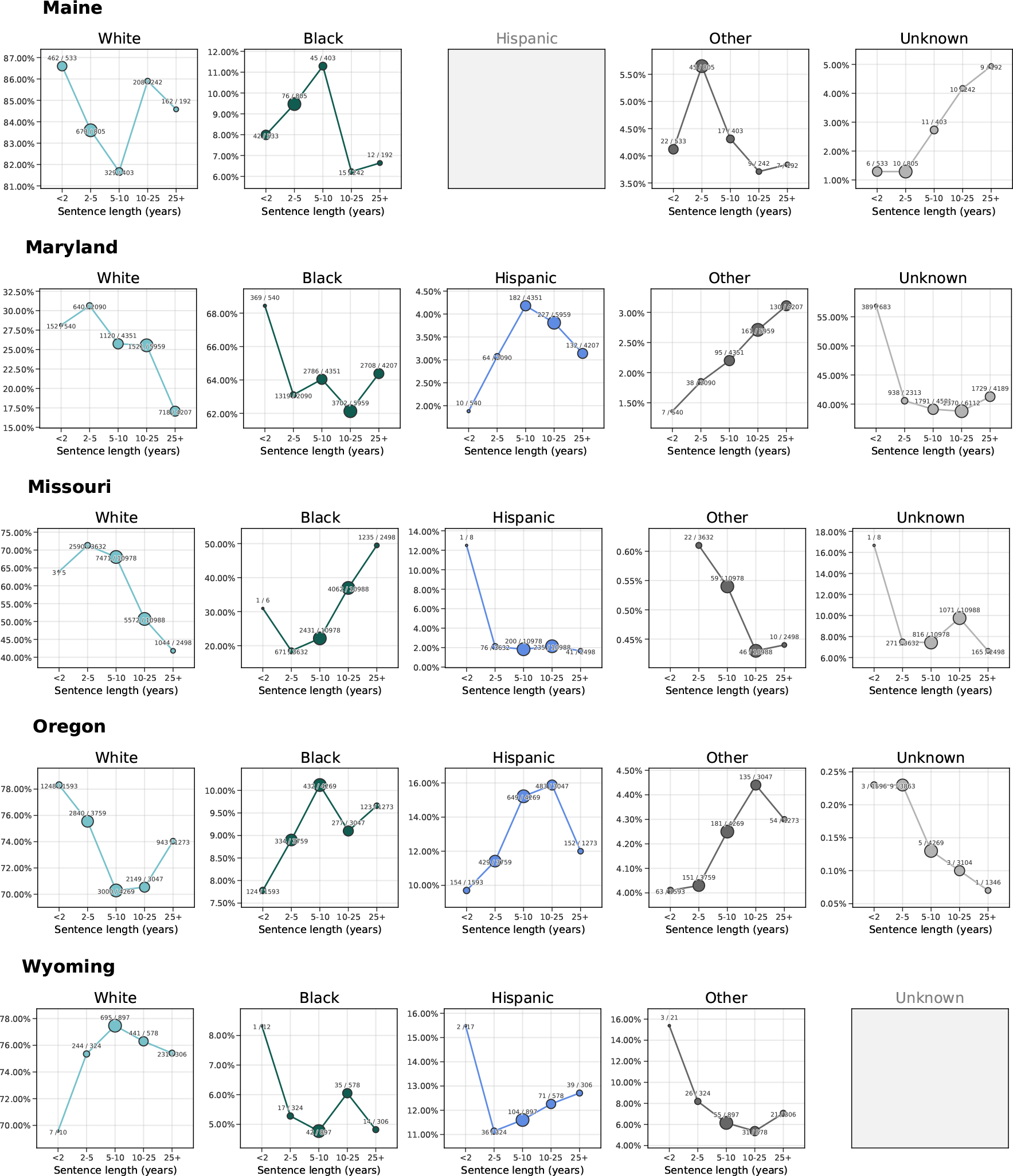
Examination of sentencing patterns in the five states that do not follow the nationwide trend from Figure 1B. **Data are from the NCRP** [1].

**Figure A.6:**
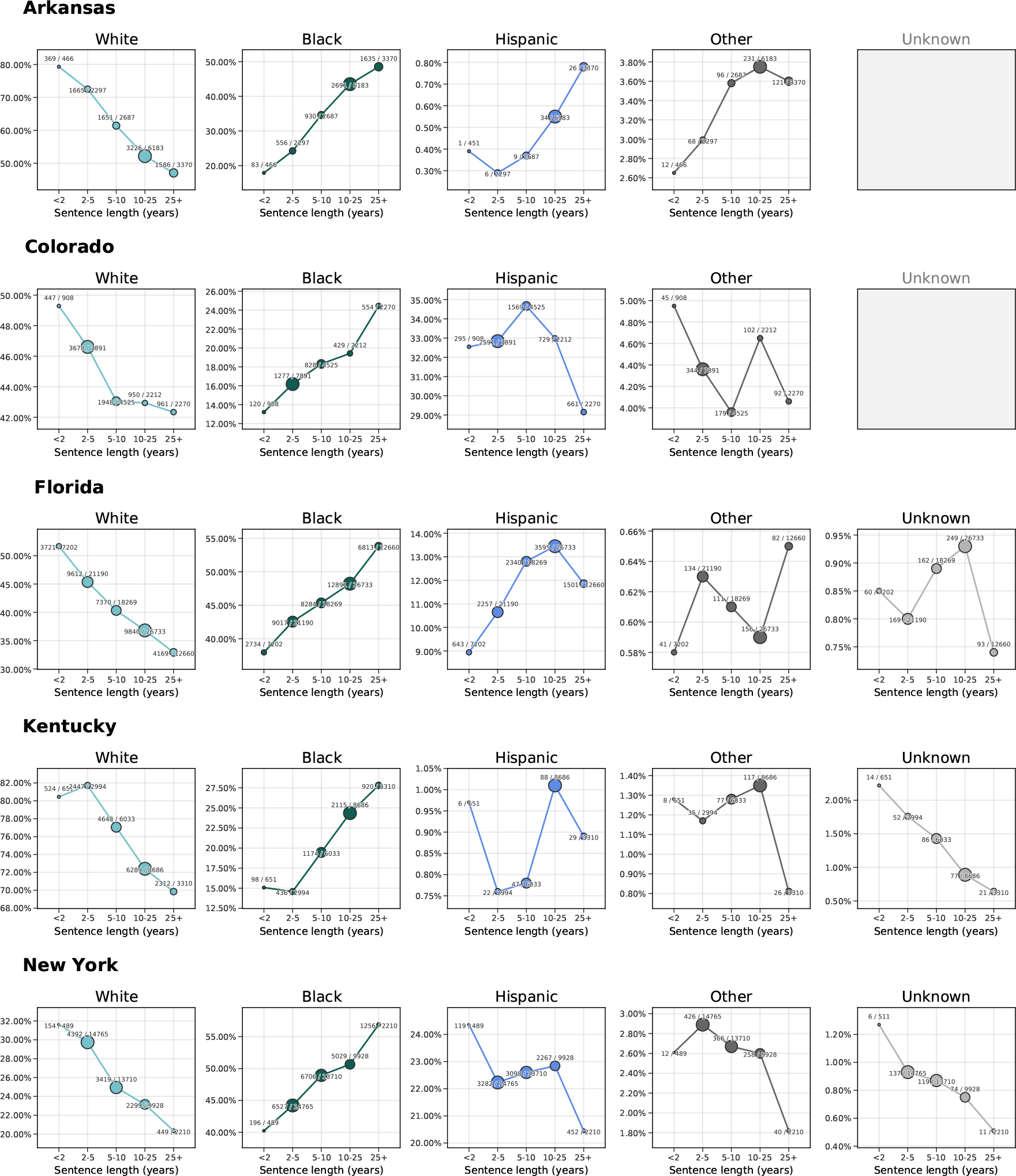
Companion figure to Figure A.5, examining sentencing patterns across five states that do follow the nationwide trend from Figure 1B. **Data are from the NCRP** [1].

**Figure A.7:**
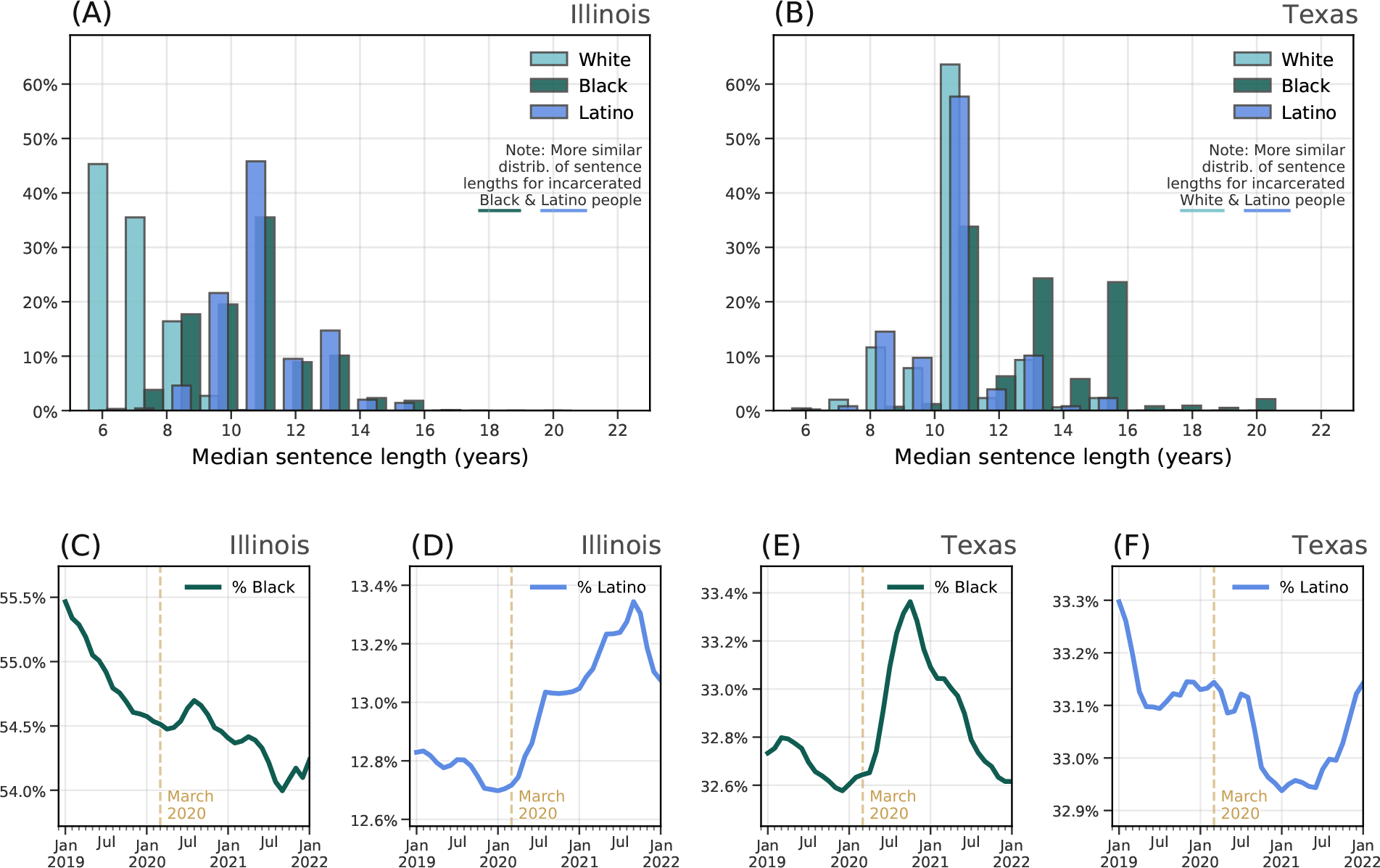
Comparison of median length of sentence among Black, White, and Latino incarcerated populations in Illinois and Texas. We randomly sample 100 incarcerated White, Black, and Latino people and report the median length of sentence; by repeating this process thousands of times, we can approximate the median length of sentence, by race in **(A)** Illinois and **(B)** Texas. For each state, we compare the percent of incarcerated people who are Black **(C & E)** and Latino **(D & F)**, highlighting the importance of racial differences in sentence length in driving the trends we observe in this study.

**Figure A.8:**
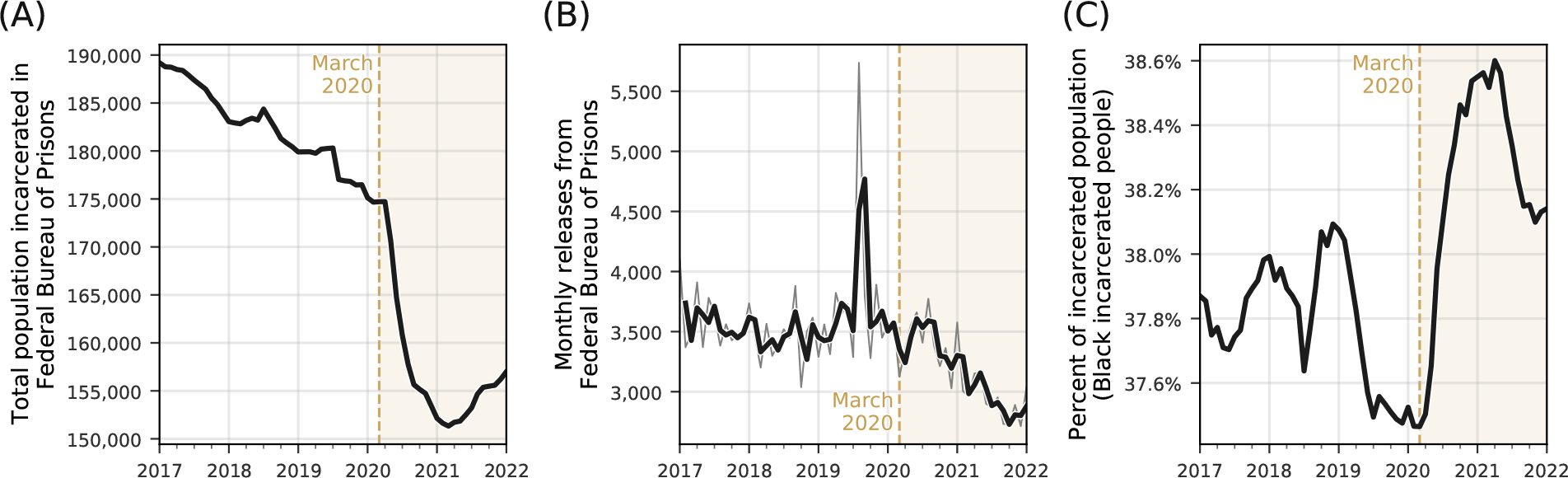
Federal Bureau of Prisons population data. (A) Total number of people incarcerated in the Federal Bureau of Prisons (FBOP) (monthly, from January 2017 until January 2022). **(B)** Total number of people released from FBOP (lighter curve: monthly; darker curve: two-month rolling average). During the pandemic, releases decreased alongside decreases in the total population; notably, we do not see months with unusually high numbers of releases once the pandemic starts. **(C)** Percent of total FBOP population who are Black.

**Figure A.9:**
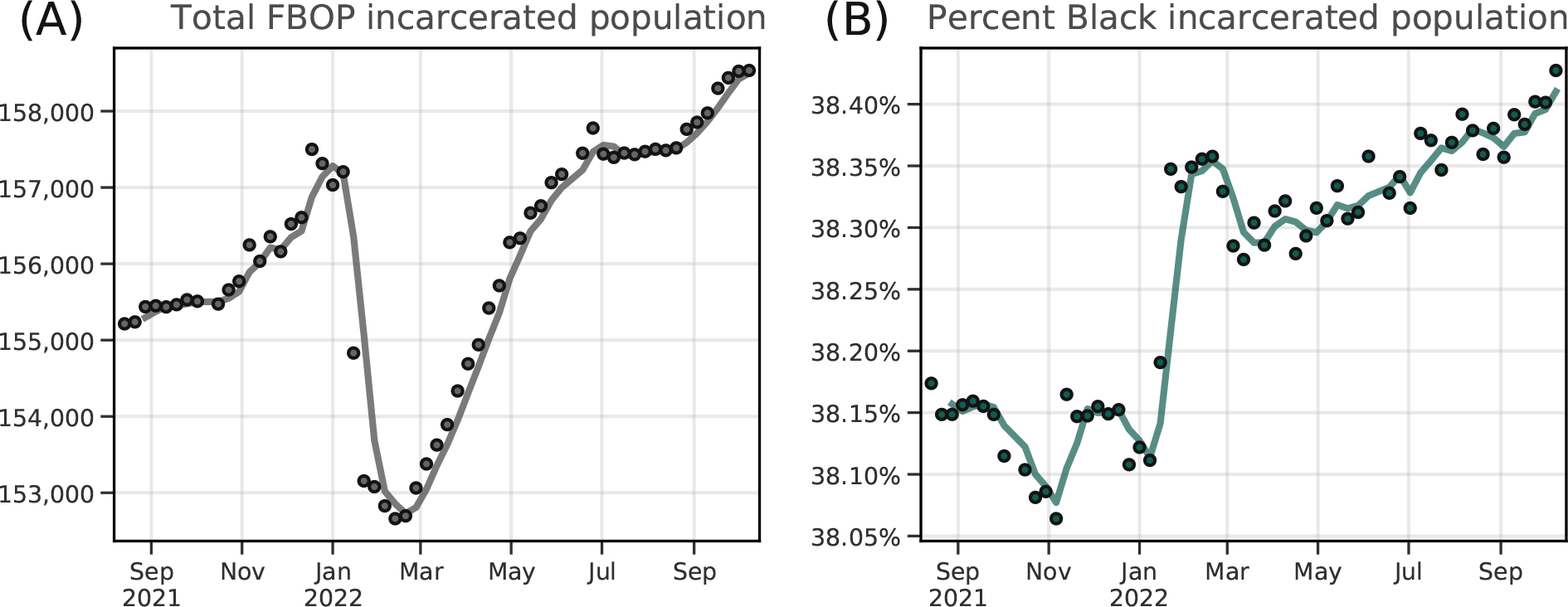
Case study: Releases and race in FBOP populations. (A) Total number of people incarcerated in the Federal Bureau of Prisons (FBOP) (weekly, from September 2021 until September 2022). **(B)** Percent of total incarcerated population who are Black.

**Figure A.10:**
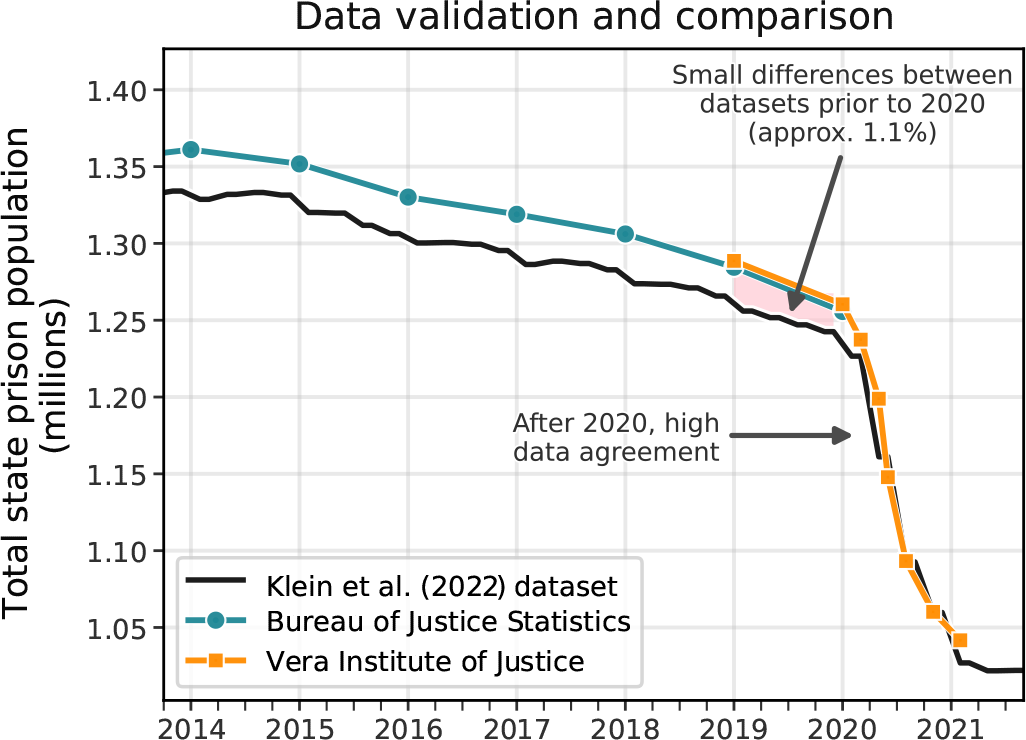
Data validation. We compare the novel data presented here to data from the Bureau of Justice Statistics [2] and the Vera Institute for Justice [3], finding high data agreement during 2020 and early 2021. There are small differences between our dataset and the Bureau of Justice Statistics prior to 2020, but see the Data & Methods section and Table for further explanation of these differences.

**Figure A.11:**
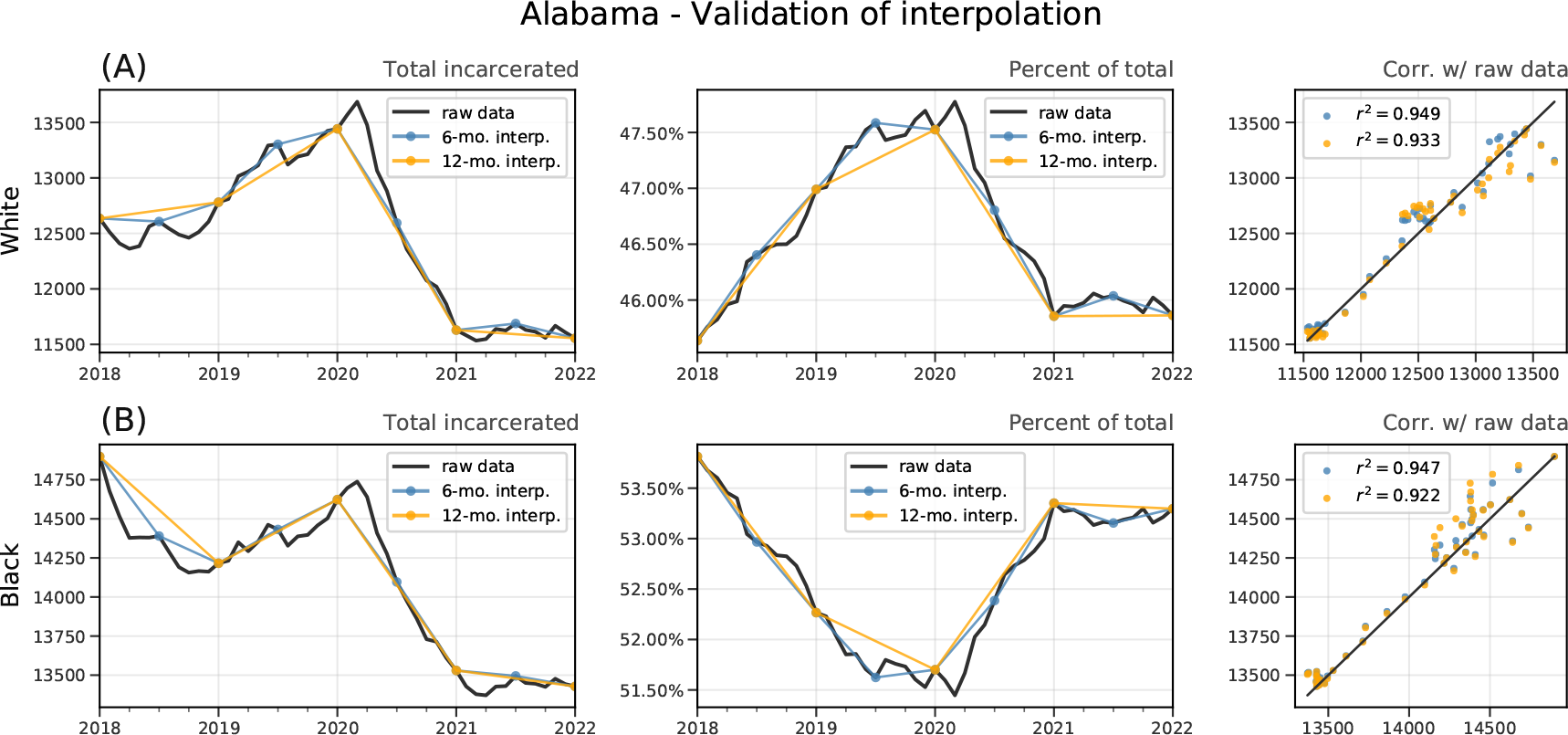
Validation of interpolation on high-quality data states: Alabama.

**Figure A.12.**
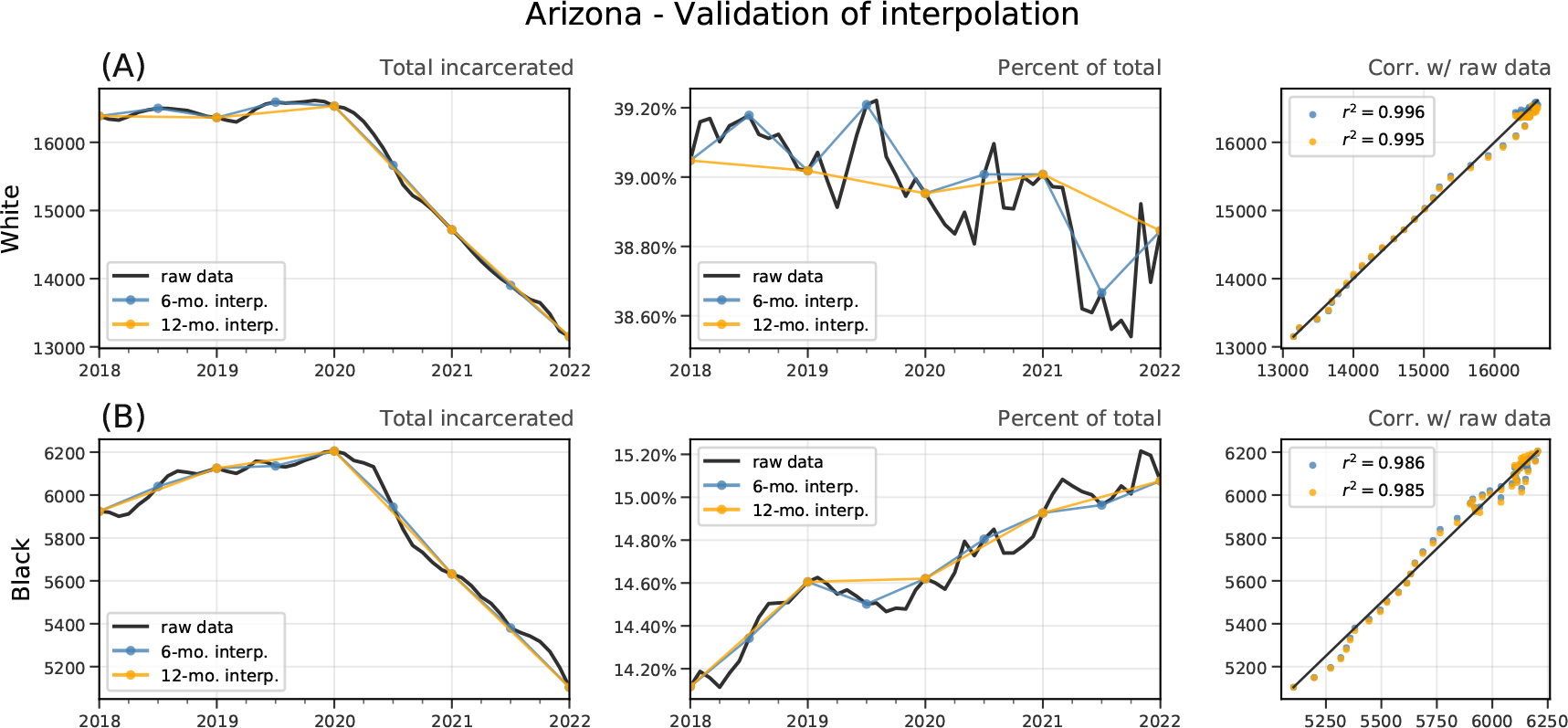
: Validation of interpolation on high-quality data states: Arizona.

**Figure A.13:**
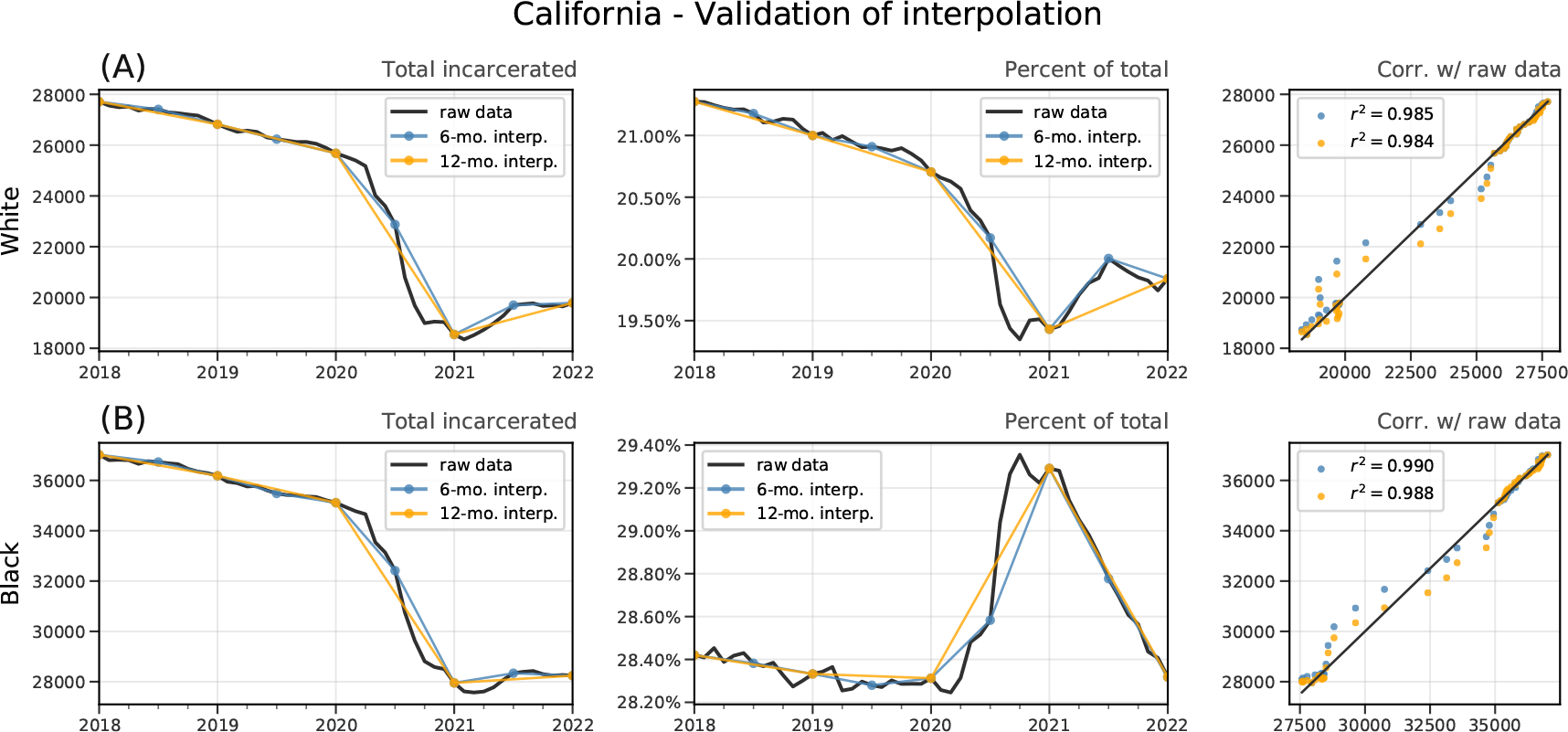
Validation of interpolation on high-quality data states: California.

**Figure A.14:**
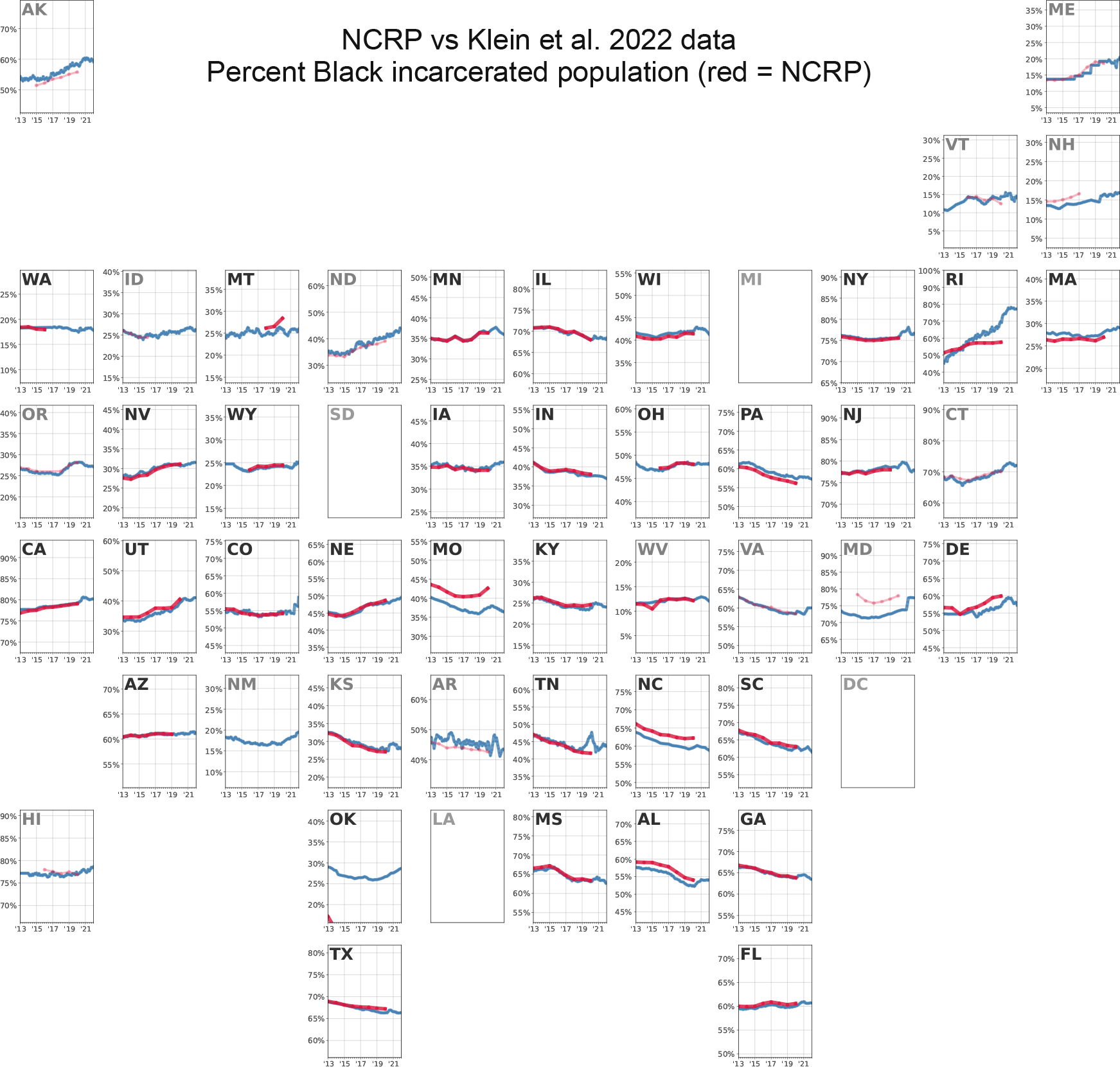
Comparison of Klein et al. dataset to historic data from the National Corrections Reporting Program data. States with light grey labels (e.g. Alaska, Oregon, Hawaii, Idaho, etc.) are not included in the “term file” (states categorized as having more reliable data). Washington, California, Nevada, Utah, Nebraska, Arizona, Colorado, Wyoming, Kansas, Texas, Iowa, Minnesota, Illinois, Indiana, Kentucky, Tennessee, Mississippi, West Virginia, Ohio, Wisconsin, Georgia, Florida, New Jersey, New York, and South Carolina are the states with the highest degree of correspondence between the two datasets.

**Figure A.15:**
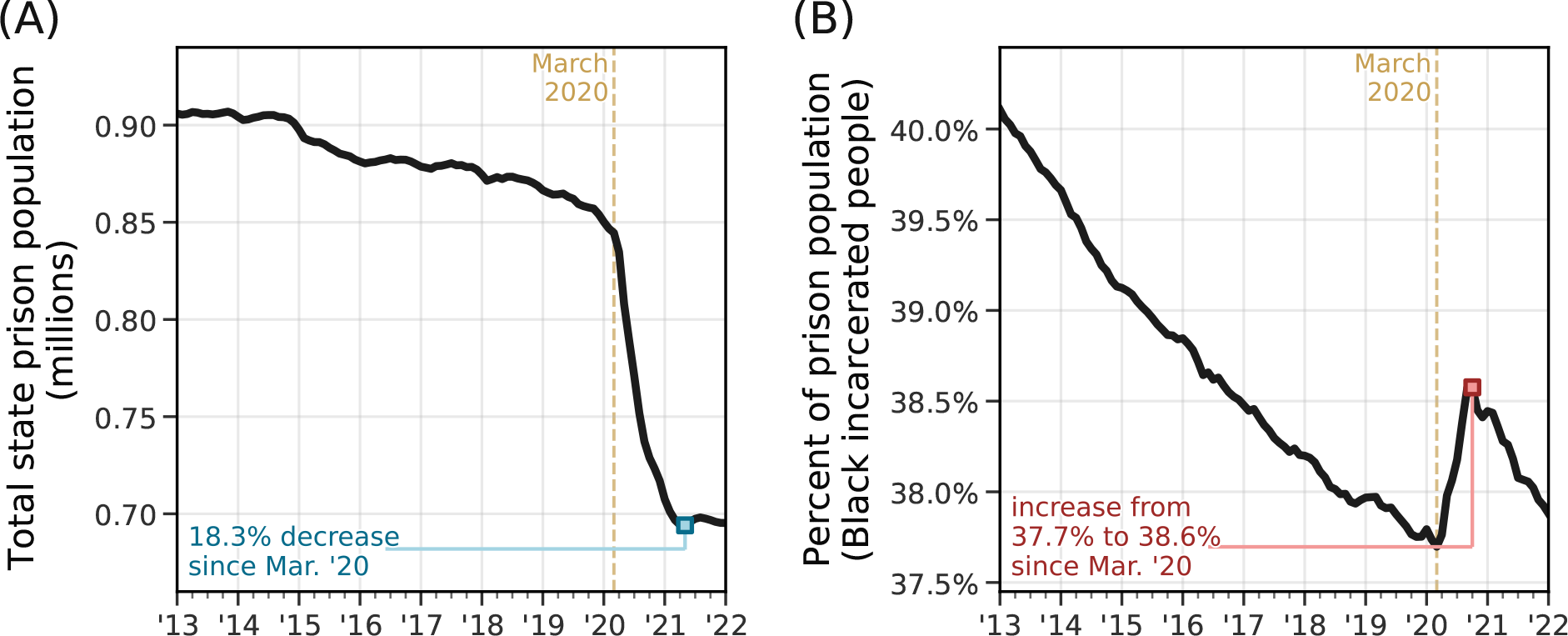
Replication of. Figure 1**, based only on data from states with high overlap with NCRP data.** If we reproduce our main result using only states with high overlap between the NCRP data and our own (a proxy for overall reporting quality: Washington, California, Nevada, Utah, Nebraska, Arizona, Colorado, Wyoming, Kansas, Texas, Iowa, Minnesota, Illinois, Indiana, Kentucky, Tennessee, Mississippi, West Virginia, Ohio, Wisconsin, Georgia, Florida, New Jersey, New York, and South Carolina), we see the same qualitative result.

**Figure A.16:**
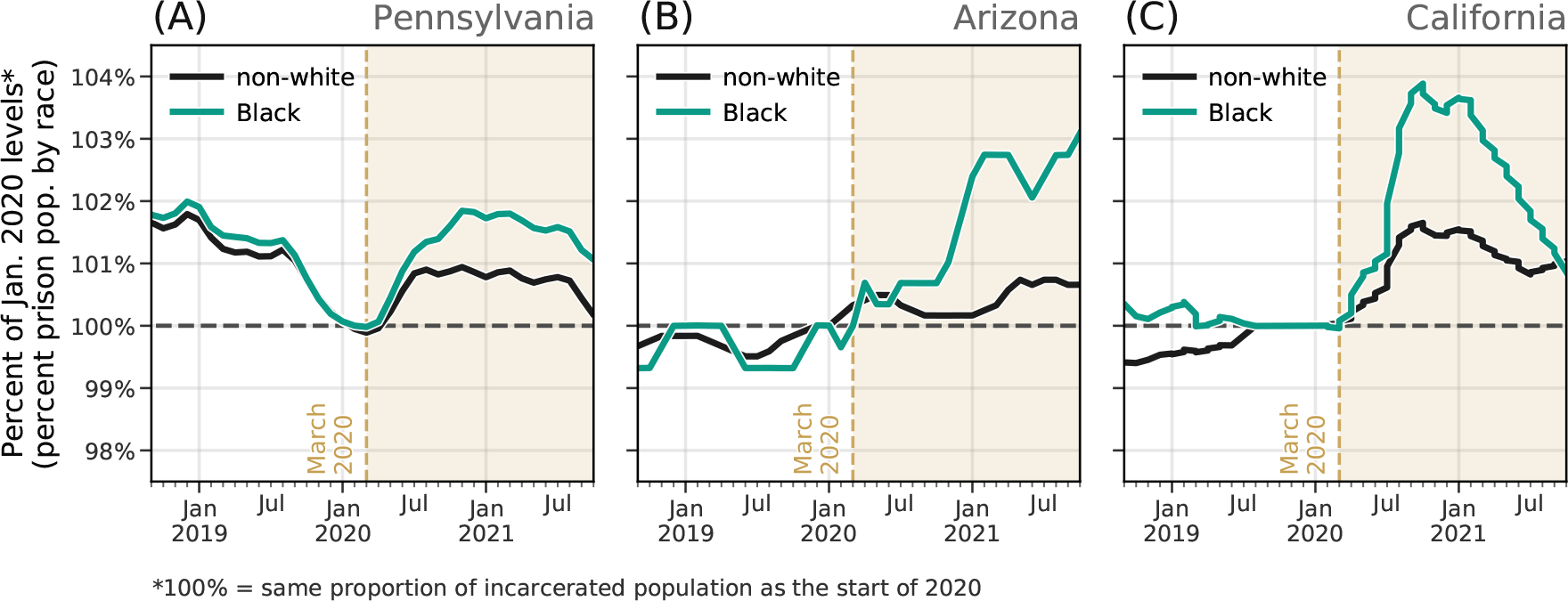
Larger effect among incarcerated Black people. Comparison of the relative increase in the proportion of incarcerated people who are non-white vs. black in three states during the pandemic. This trend is especially pronounced in the three states above, however there are several states where the opposite is true. Further distinguishing these effects will be the subject of future research.

**Figure A.17:**
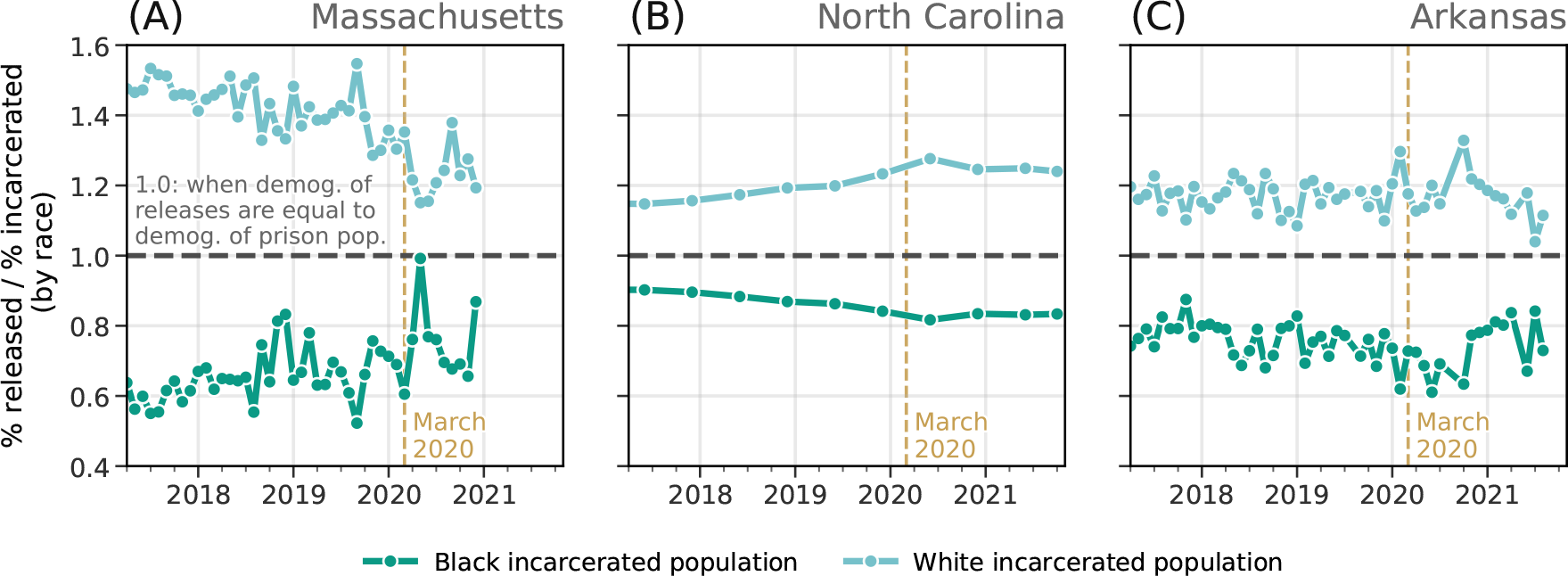
Ratio of race of releases to race of prison population. Here, we highlight the evolution of the ratio between *r_x_/N_x_*, where *r_x_* is the percent of releases who are race *x* and *N_x_* is the percent of the incarcerated population who are race *x* (in this case, we use Black and white incarcerated people for *x*). If this ratio is 1.0, there is a proportional number of releases as one would expect, given the demographic composition of the prison population. In the three states included here, white incarcerated people account for a larger share of releases than one would expect, given the demographic distribution of the prison population; conversely, incarcerated Black people are released at lower-than-expected rates.

**Figure A.18:**
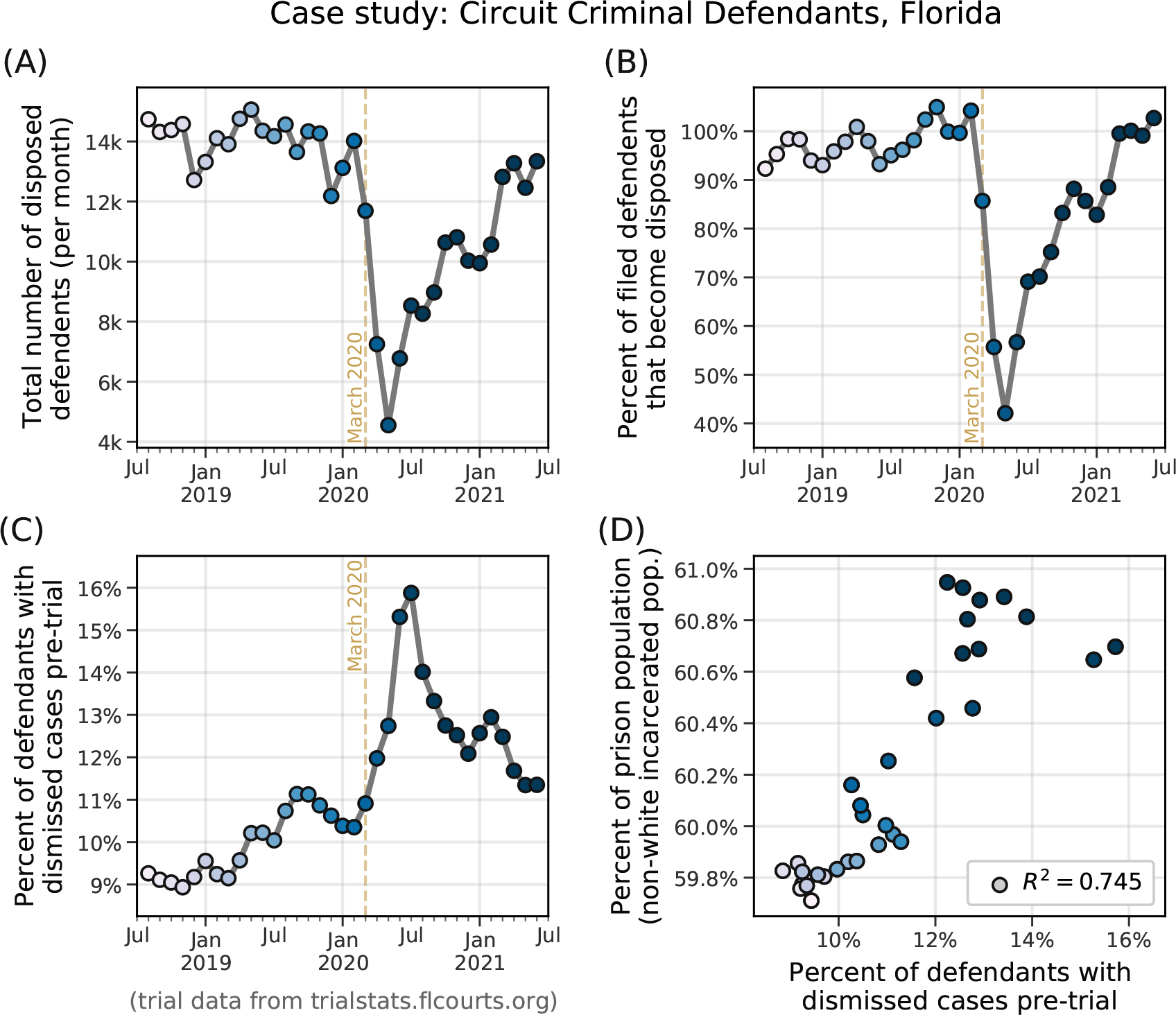
Circuit criminal defendants in Florida. (A) Total number of defendants with disposed cases (i.e., number of closed cases) over time. **(B)** Case completion rate (number of disposed defendants divided by the number of cases filed). **(C)** Percent of defendants with cases that are dismissed before going to trial (different from pre-trial guilty pleas). See Figure A.19 for preliminary data about the demographics of defendants with dismissed cases—namely, that there is a relative increase in white defendants with dismissed cases during this period. **(D)** Correlation between percent of dismissed cases and percent of incarcerated individuals who are non-white (one month lag). Data from Florida Office of the State Courts Administrator [19], from July 30, 2018 to June 30, 2021 (latest data available).

**Figure A.19:**
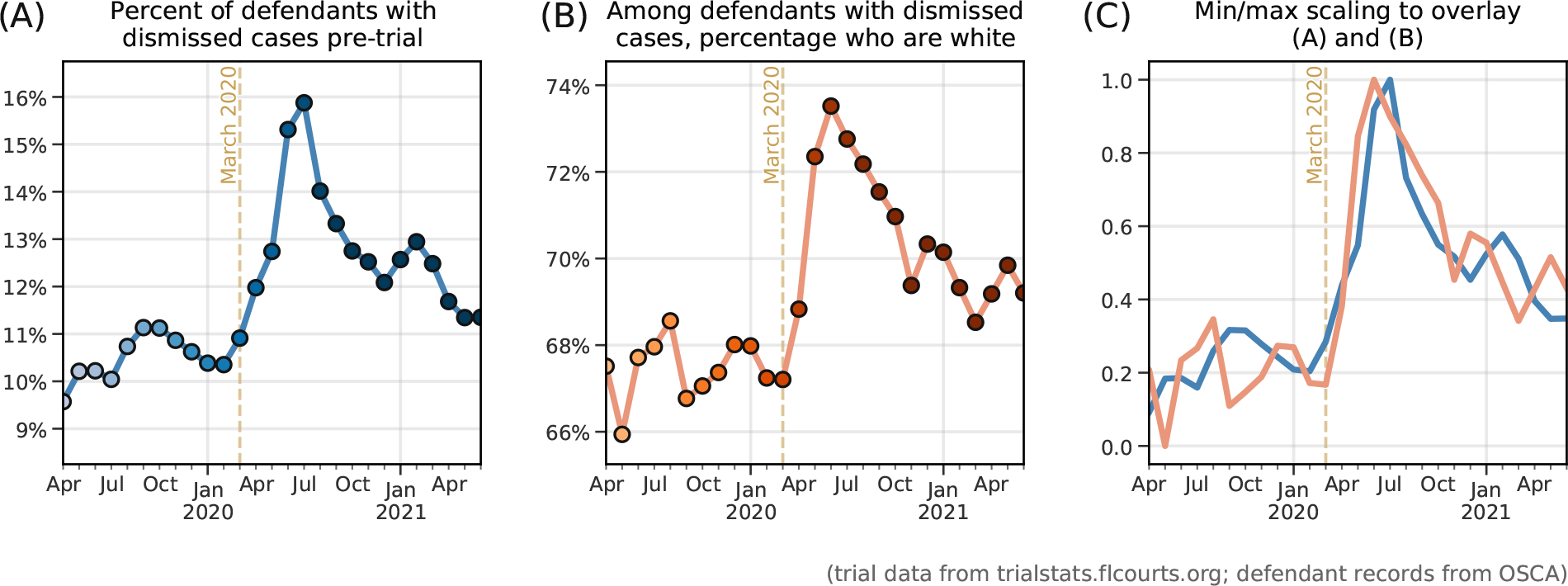
Florida pre-trial court dismissals, by race. (A) Reproduction of Figure A.18C. **(B)** Using data compiled by the Florida Office of the State Courts Administrator (OSCA) from the Criminal Transaction System (CTS), we can begin to get a sense of the demographics of the defendants with dismissed cases (Note: OSCA stresses that these data are provided by court clerks and are subject to change, and that the conclusions and analysis that is done using this data are solely those of the authors of this paper). **(C)** Comparison of the curves in **(A)** and **(B)** via min-max scaling.

**Figure A.20:**
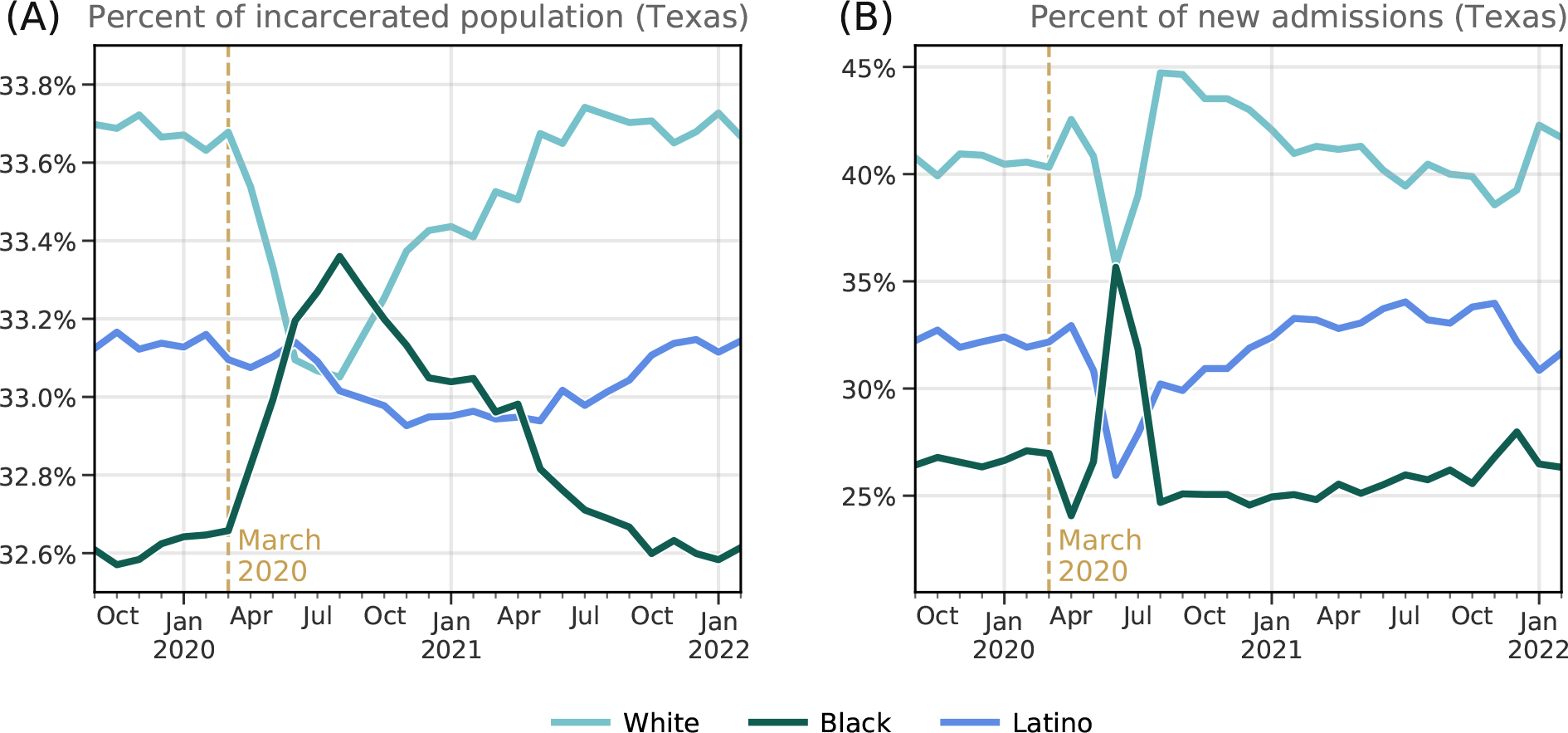
Race of incarcerated population and monthly admissions in Texas. Incarcerated population in Texas over time, by race. (B) Monthly percentage of new admissions into Texas state prisons, by race.

**Figure A.21:**
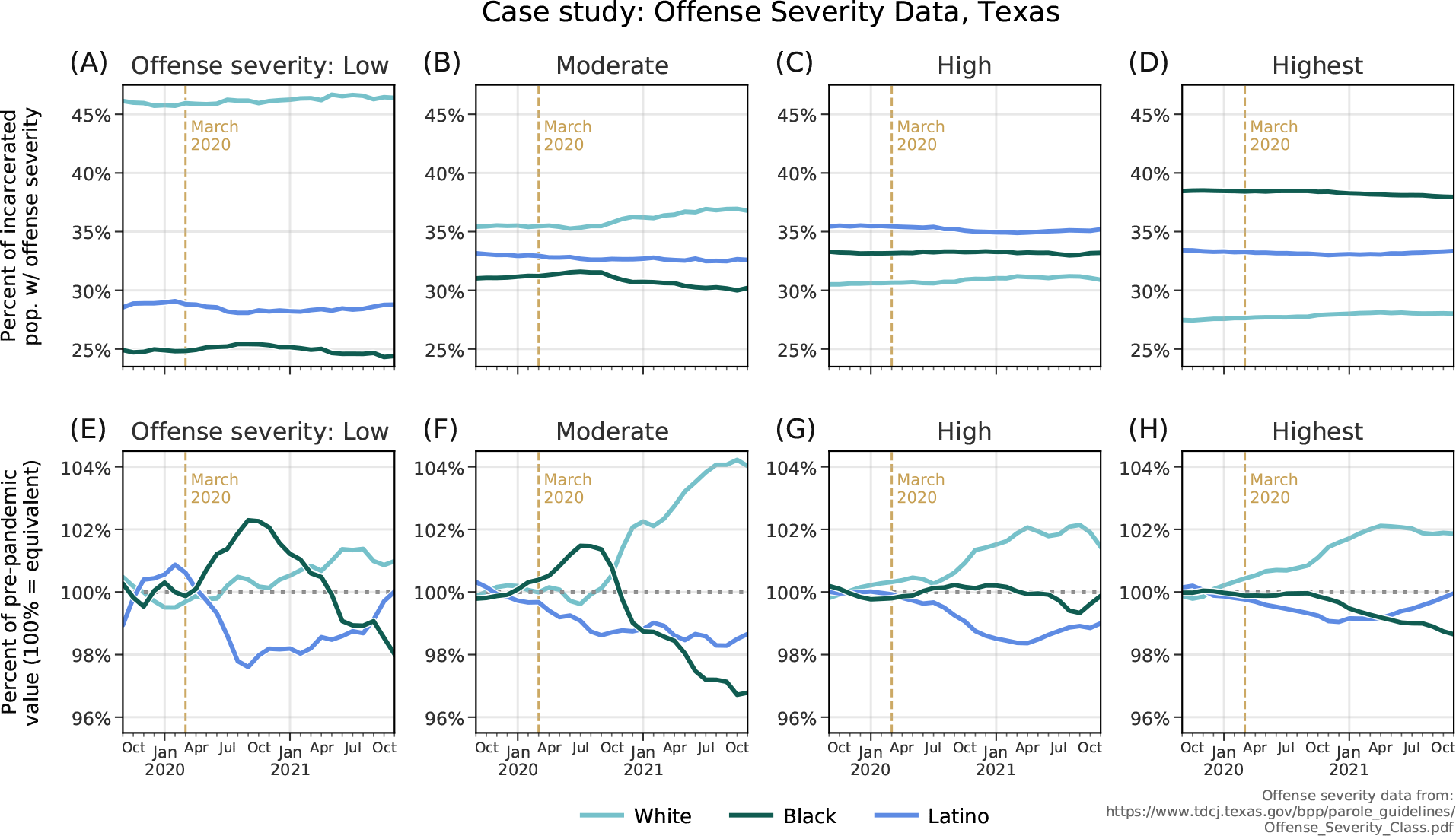
Offense severity of incarcerated persons in Texas, by race. Top row (A-D): For each offense-severity category (low, moderate, high, and highest), we plot monthly time series of the percent of each group who are White, Black, or Latino incarcerated people. Bottom row (E-H): The same curves as A-D, standardized by each race group’s average values prior to March 2020. A value of 100% indicates no change relative to pre-pandemic averages.

**Figure A.22:**
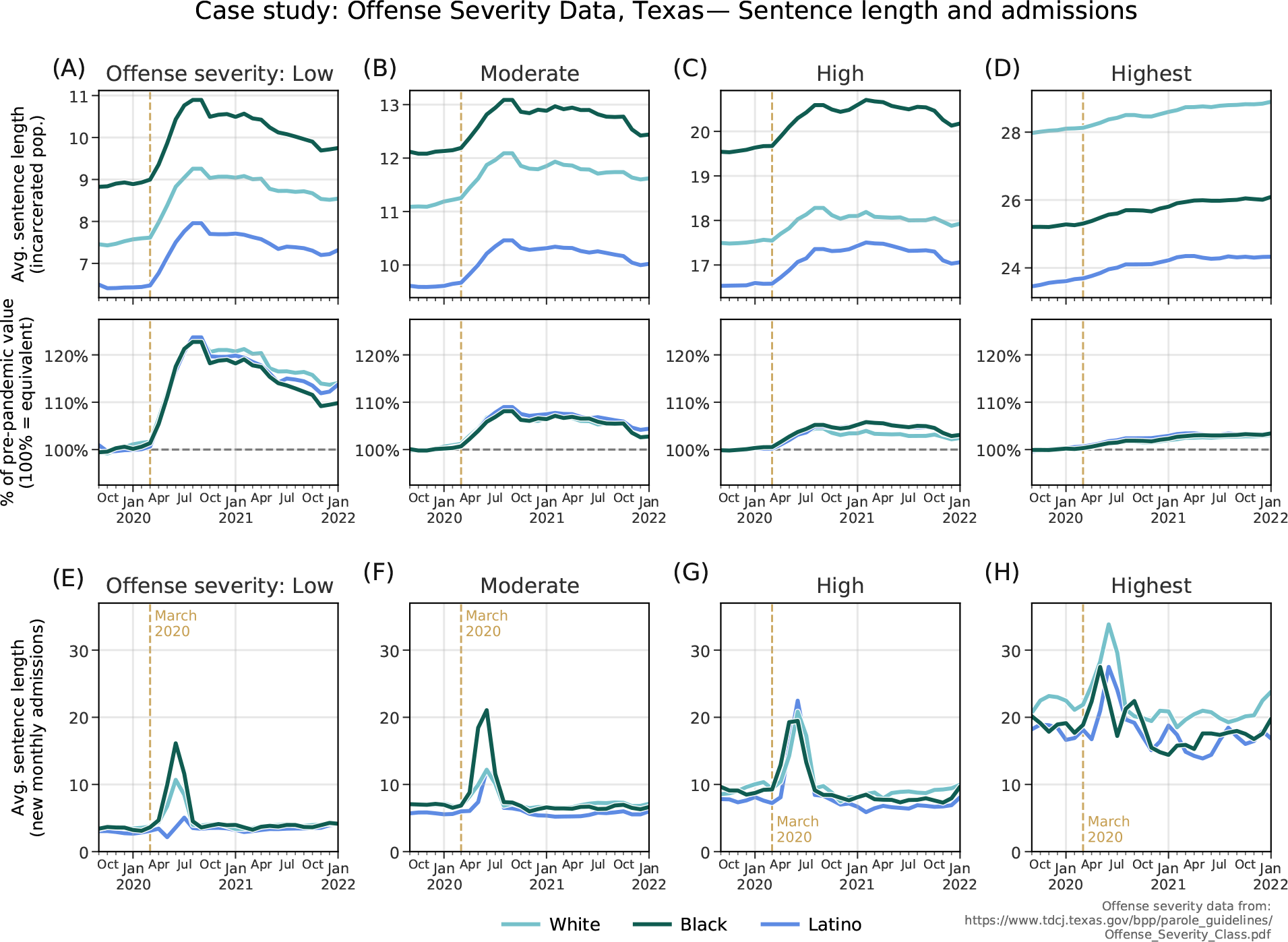
Comparison of average sentence length in Texas, by race and offense severity. (A-D) Top: Average sentence length of incarcerated population in Texas state prisons (excluding life-sentences and sentences over 80 years), by race and offense severity. Bottom: Change relative to pre-pandemic values (100% indicates no change). For all offense types, we see increases in the average sentence length at the start of the pandemic due to the drop in monthly prison admissions without commensurate decreases in releases. **(E-H)** Average sentence length of new prison admissions, by offense severity and race.

**Figure A.23:**
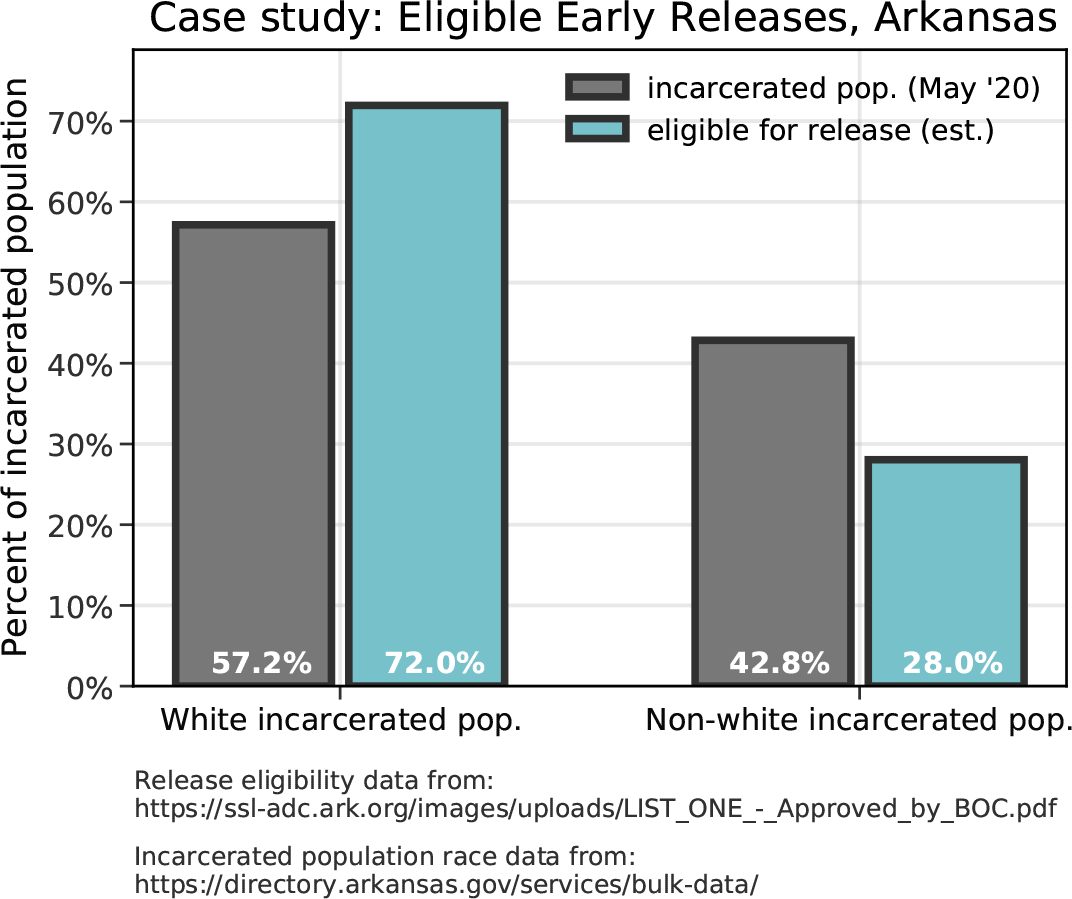
Case study: Eligible releases in Arkansas. In May 2020, incarcerated white people accounted for 57.2% of the total prison population in Arkansas. However, white incarcerated people accounted for over 72% of the those listed as eligible for early release under public health precautions for COVID-19 [24].

**Table A.1:** **Overview of Klein et al. dataset.** This dataset includes prison population time series for all 50 states and the District of Columbia. Here, we have included hyperlinks to each state’s website where we started to collect the data. For some states, we submitted public records requests in order to obtain data about inmate race statistics.

| State | Primary source of data | Frequency | Data start | Data end |
| --- | --- | --- | --- | --- |
| Alabama | Inmate Search website | weekly | Sep. 2000 | Jan. 2022 |
| Alaska | Research Records website | monthly | Jan. 2015 | Jan. 2022 |
| Arizona | Corrections at a Glance website | monthly | Jan. 2017 | Jan. 2022 |
| Arkansas | Director's Board Reports | monthly | Jan. 2000 | Jan. 2022 |
| California | Offender Population Reports and public records request | monthly | Jan. 2014 | Jan. 2022 |
| Colorado | Inmate Population Profile | monthly | Jun. 1992 | Jan. 2022 |
| Connecticut | Monthly Statistics | monthly | Jan. 2010 | Jan. 2022 |
| Delaware | Inmate Population Data | monthly | Jan. 2017 | Jan. 2022 |
| Washington D.C. | Inmate Demographics and Statistics | quarterly | Oct. 2015 | Jan. 2022 |
| Federal BOP | Inmate Race Statistics and public records request | weekly | Jan. 2011 | Jan. 2022 |
| Florida | Statistics and Publications and public records request | monthly | Jan. 2005 | Jan. 2022 |
| Georgia | Monthly Profile of All Inmates | monthly | Jan. 2006 | Jan. 2022 |
| Hawaii | Annual Reports and public records request | monthly | Mar. 2018 | Jan. 2022 |
| Idaho | Incarcerated Population Report and public records request | monthly | Jan. 2013 | Jan. 2022 |
| Illinois | Prison Population Data Sets | quarterly | Jun. 2005 | Jan. 2022 |
| Indiana | Fact Cards | quarterly | Jan. 2002 | Jan. 2022 |
| Iowa | Quarterly Quick Facts and public records request | monthly | Feb. 2005 | Jan. 2022 |
| Kansas | Population Reports | monthly | Jun. 1990 | Jan. 2022 |
| Kentucky | Monthly Reports | monthly | Dec. 2011 | Jan. 2022 |
| Louisiana | Annual Statistics and public records request | 2x-yearly | Jan. 1989 | Jan. 2022 |
| Maine | Reports & Statistical Data | monthly | Jan. 2015 | Jan. 2022 |
| Maryland | Inmate Characteristics Reports and public records request | quarterly | Jul. 1990 | Jan. 2022 |
| Massachusetts | Institutional Fact Cards and Research Reports | monthly | Jan. 2012 | Jan. 2022 |
| Michigan | Statistical Reports | yearly | Jan. 1961 | Jan. 2022 |
| Minnesota | Historical Population Summary Reports | 2x-yearly | Jan. 1998 | Jan. 2022 |
| Mississippi | Monthly Fact Sheets | monthly | Apr. 2001 | Jan. 2022 |
| Missouri | Publications and Information & Missouri Sunshine Law | monthly | Jun. 2004 | Jan. 2022 |
| Montana | Data & Statistics and public records request | monthly | Jan. 2005 | Jan. 2022 |
| Nebraska | Statistical Reports | quarterly | Dec. 2007 | Jan. 2022 |
| Nevada | Weekly Fact Sheets | weekly | Jan. 2015 | Jan. 2022 |
| New Hampshire | NHDOC Reports | monthly | Jul. 1999 | Jan. 2022 |
| New Jersey | Offender Stats | yearly | Jan. 2011 | Jan. 2022 |
| New Mexico | Notice & Reports and public records request | monthly | Jan. 2010 | Jan. 2022 |
| New York | Inmates Under Custody and public records request | monthly | Jan. 2000 | Jan. 2022 |
| North Carolina | DPS Research & Planning | quarterly | Jan. 1996 | Jan. 2022 |
| North Dakota | Data Narratives: Prison | monthly | Nov. 2001 | Jan. 2022 |
| Ohio | Monthly Fact Sheets | monthly | Aug. 2016 | Jan. 2022 |
| Oklahoma | Offender Information and public records request | weekly | Jul. 2010 | Jan. 2022 |
| Oregon | Inmate Population Profile and public records request | monthly | Jan. 1998 | Jan. 2022 |
| Pennsylvania | Monthly Population Reports and public records request | monthly | Feb. 2017 | Jan. 2022 |
| Rhode Island | Offender Statistics & Reports and public records request | monthly | Jan. 2012 | Jan. 2022 |
| South Carolina | Archived SCDC FAQs | monthly | Jul. 2011 | Jan. 2022 |
| South Dakota | Adult Inmates by Race/Ethnicity | monthly | Dec. 2016 | Jan. 2022 |
| Tennessee | Felon Population Reports | monthly | Jul. 2004 | Jan. 2022 |
| Texas | Inmate Population High Value Dataset and public records request | monthly | Sep. 2005 | Jan. 2022 |
| Utah | UDC Statistics and public records request | monthly | Sep. 2012 | Jan. 2022 |
| Vermont | Population Report | monthly | Jan. 2020 | Jan. 2022 |
| Virginia | Offender Population Reports | monthly | Jul. 2010 | Jan. 2022 |
| Washington | Statistical Reports | quarterly | Nov. 2009 | Jan. 2022 |
| West Virginia | Adult prison statistics and public records request | 2x-yearly | Jul. 2002 | Jan. 2022 |
| Wisconsin | Data & Reports | monthly | Jul. 1990 | Jan. 2022 |
| Wyoming | Population Demographics Report and public records request | quarterly | Jan. 2006 | Jan. 2022 |

**Table A.2:**
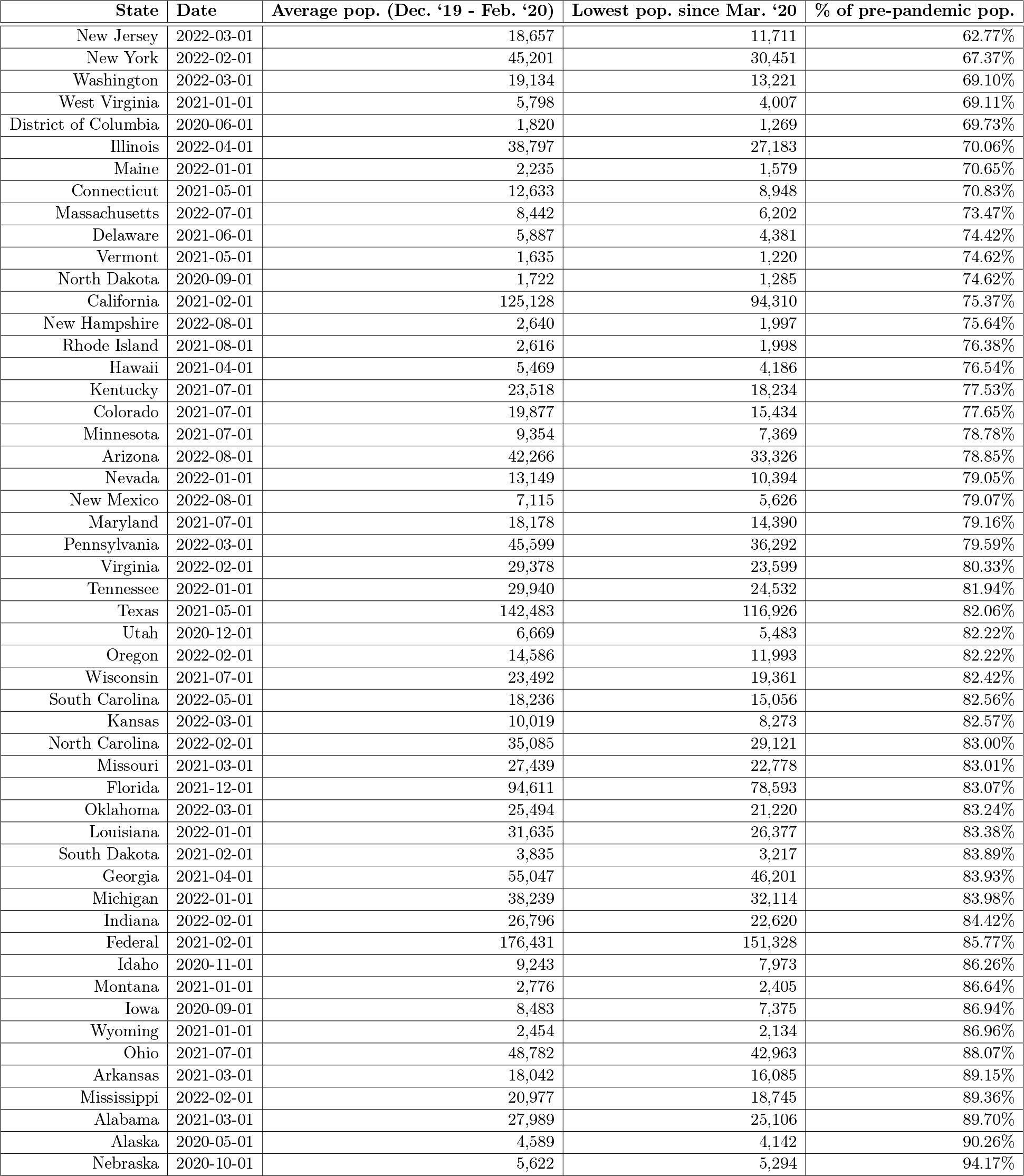
**Extent of prison population reductions.** For each state, we list the date (after March 2020) that the prison population reached its lowest reported value, as well as the magnitude of each state’s reduction (measured as the percent of pre-pandemic prison population). States are listed in ascending order by largest population reductions.

**Table A.3:** **Five states with the largest differences to BJS data.** Because there were slight discrepancies between the data collected in the current study and the yearly data released by the BJS, we include this table, which links to the data sources used. These data were collected from states’ Departments of Corrections websites (or obtained through Freedom of Information Act requests). For both of these states, the BJS includes a disclaimer. Montana: “Data for 2019 are not comparable to data for previous years.” Ohio: “Includes a small number of incarcerated individuals sentenced to one year or less.” For Texas and Virginia, we include the link to directly access data reported by the state for the time period in question, and in each case, the BJS data does not correspond to data reported by the state. The discrepancies in Florida’s numbers are relatively small compared to the overall number of people in prison. As stated above, we are confident in the data collection here, which involved making successful public records requests to the Department of Corrections.

| State | Source | BJS: State Prison Pop.<br>(as of Dec. 31, '19) | Klein et al.: State Prison<br>Pop. (as of Dec. 31, '19) | Difference |
| --- | --- | --- | --- | --- |
| Texas | Demographic Highlights, On Hand [link] | 158,429 | 142,169 | 16,260 |
| Virginia | Monthly Population Summary [link] | 36,091 | 29,233 | 6,858 |
| Montana | Public records request; [link] | 4,723 | 2,806 | 1,917 |
| Florida | Public records request; [link] | 96,009 | 94,197 | 1,812 |
| Ohio | Monthly Fact Sheet [link] | 50,338 | 48,697 | 1,641 |

**Table A.4:** **Method of reporting race of incarcerated persons, by state**. As of May 15, 2022, we have yet to receive confirmation from Alabama, District of Columbia, and Louisiana about their policies, but for nearly every state in this table, we see an explicit policy of recording racial categories via self-report.

| State | Method | Source | Relevant language |
| --- | --- | --- | --- |
| AL | × | × | × |
| AK | self-report | emailed | Race is self-identified by the inmate. |
| AZ | self-report | emailed | The data is self-reported/self-identifying. |
| AR | self-report | emailed | During the intake process, individuals self-report their race to staff. |
| CA | self-report | document | Ethnicity is self-reported by offenders who choose from a list of 28 ethnicity types. |
| CO | self-report | document | Data from the DOC were used... because it contained self-reported ethnicity for offenders. |
| CT | self-report | phone | Returned phone call to confirm the policy is self-report. |
| DE | self-report<sup>‡</sup> | document | e.g. “Delaware’s Level IV population identified as...” |
| DC | × | × | × |
| FBOP | self-report | document | Race/ethnic categories are presented as separate... individuals can select multiple racial categories. |
| FL | self-report | phone | Returned phone call to confirm the policy is self-report. |
| GA | self-report | emailed | The data is self-reported at admission. |
| HA | self-report | emailed | Ethnicity is self-reported; PSD does not verify the accuracy of the information given. |
| ID | self-report | emailed | It is self-report within the pre-sentence investigation process. |
| IL | self-report | emailed | Self-report. IDOC does not verify the information. |
| IN | self-report | emailed | The offenders self-identify at the time of entry into our pre-determined categories (not free form). |
| IA | self-report | emailed | We collect from inmate self-identifying and court documents. |
| KS | self-report | emailed | Demographic info, including race, is collected as part of the intake process. Individuals self-identify. |
| KY | self-report | phone | Returned phone call to confirm the policy is self-report. |
| LA | × | × | × |
| ME | self-report | emailed | This information is self-reported by the resident during intake. |
| MD | self-report* | emailed | Both self-report and staff-assigned. |
| MA | self-report* | phone | It is mostly self-reported, but if the county sends a face sheet the Booking Officer will use that. |
| MI | self-report | emailed | Each inmate self-identifies to staff for the sake of collecting data that is used in statistical reports. |
| MN | self-report* | document | Race information may be self-identified or classified by an observer. |
| MS | self-report | emailed | Race is self-reported during the intake process. |
| MO | self-report | emailed | The offender is self-identifying. |
| MT | self-report | emailed | Self-reported, unless the individual refuses... [then] race would be estimated by staff during intake. |
| NE | self-report | emailed | The race data that we use for our statistical reports is self-reported by the inmate at intake. |
| NV | self-report<sup>‡</sup> | document | Demographic information is collected during intake... data are gathered from inmates progressively. |
| NH | self-report | emailed | The residents do self report upon intake. Thank you for your inquiry and have a great day. |
| NJ | self-report | emailed | Race and ethnicity are verified through self-report at intake. |
| NM | self-report | emailed | The inmates self-identify. |
| NY | self-report | document | The information about racial and ethnic origin is self-reported. |
| NC | self-report | emailed | Race is self-identified via a questionnaire that is administered during the entrance interview. |
| ND | self-report | emailed | Race information is self-reported. |
| OH | self-report<sup>‡</sup> | document | Information collected from incarcerated individuals: Race/ethnic origin, nationality, age... |
| OK | self-report | emailed | Yes, it’s self-report by the inmate. |
| OR | self-report<sup>‡</sup> | document | HB 3064, passed August 2019, effective January 2020 |
| PA | self-report | emailed | The PA DOC racial data is self-reported by the inmate at intake. |
| RI | self-report | document | ...we cannot determine whether inmates identifying themselves as Hispanic are white or black. |
| SC | self-report | document | Based on inmate self-reported information at intake. |
| SD | self-report | document | ...collect specified demographic information (religious preference, age, race, height, weight, etc.). |
| TN | self-report | emailed | The information is self-reported. |
| TX | visual | emailed | ...during intake, the TDCJ will visually determine the race of the individual. |
| UT | self-report | emailed | Race is inmate self-identifying. |
| VT | self-report | document | Some categories are combined due to the low number of individuals who identify with these races... |
| VA | self-report | emailed | Virginia collects race information from several different sources, as well as being self-reported. |
| WA | self-report | document | Race and ethnicity are self-reported and grouped to include the following... |
| WV | self-report | emailed | No problem, in WV race is based on inmates self identification at intake. |
| WI | self-report | emailed | When an individual enters prison they are asked which race designation best describes them. |
| WY | self-report | phone | Returned phone call to confirm the policy is self-report. |

\* = Nonspecific policy; <sup>‡</sup> = Documentation is suggestive of self-report; × = could not confirm data.

**Table A.5:** **Statewide statistics for Florida Circuit Criminal Defendants.** Data collected from the Trial Court Statistics Search (http://trialstats.flcourts.org/). Note the substantial drop-off in April and May of 2020 (highlighted above), corresponding to the period when Florida courts were closed or operating at highly reduced capacities.

| Date | Total Defendants Filed | Total Disposed | Total Disposed Pre-trial | Pre-trial: Total Dismissed | Pre-trial: Total Guilty Plea | Pre-trial: Other |
| --- | --- | --- | --- | --- | --- | --- |
| 2017-07-01 | 14,883 | 12,442 | 12,199 | 1,094 | 9,604 | 1,501 |
| 2017-08-01 | 16,621 | 16,362 | 15,885 | 1,321 | 12,838 | 1,726 |
| 2017-09-01 | 12,109 | 10,461 | 10,148 | 913 | 8,043 | 1,192 |
| 2017-10-01 | 15,297 | 15,674 | 15,180 | 1,350 | 12,276 | 1,554 |
| 2017-11-01 | 13,848 | 13,799 | 13,403 | 1,131 | 10,748 | 1,524 |
| 2017-12-01 | 13,217 | 11,267 | 10,961 | 1,053 | 8,623 | 1,285 |
| 2018-01-01 | 15,752 | 14,322 | 13,845 | 1,408 | 10,908 | 1,529 |
| 2018-02-01 | 14,156 | 13,628 | 13,187 | 1,157 | 10,499 | 1,531 |
| 2018-03-01 | 15,309 | 13,956 | 13,579 | 1,145 | 10,837 | 1,597 |
| 2018-04-01 | 15,167 | 13,757 | 13,368 | 1,097 | 10,760 | 1,511 |
| 2018-05-01 | 15,590 | 14,643 | 14,170 | 1,228 | 11,332 | 1,610 |
| 2018-06-01 | 15,470 | 14,509 | 14,097 | 1,199 | 11,119 | 1,779 |
| 2018-07-01 | 15,265 | 14,080 | 13,874 | 1,324 | 11,150 | 1,400 |
| 2018-08-01 | 16,667 | 15,399 | 15,146 | 1,405 | 12,209 | 1,532 |
| 2018-09-01 | 13,475 | 13,233 | 13,046 | 1,204 | 10,488 | 1,354 |
| 2018-10-01 | 15,749 | 15,530 | 15,299 | 1,397 | 12,510 | 1,392 |
| 2018-11-01 | 13,904 | 13,634 | 13,433 | 1,210 | 10,744 | 1,479 |
| 2018-12-01 | 13,124 | 11,798 | 11,602 | 1,118 | 9,128 | 1,356 |
| 2019-01-01 | 15,431 | 14,839 | 14,665 | 1,430 | 11,444 | 1,791 |
| 2019-02-01 | 13,997 | 13,376 | 13,175 | 1,184 | 10,357 | 1,634 |
| 2019-03-01 | 14,423 | 14,440 | 14,237 | 1,364 | 10,990 | 1,883 |
| 2019-04-01 | 14,823 | 15,069 | 14,851 | 1,461 | 11,441 | 1,949 |
| 2019-05-01 | 15,973 | 15,054 | 14,627 | 1,615 | 11,052 | 1,960 |
| 2019-06-01 | 14,810 | 13,659 | 13,281 | 1,327 | 9,781 | 2,173 |
| 2019-07-01 | 15,004 | 14,689 | 14,311 | 1,523 | 11,340 | 1,448 |
| 2019-08-01 | 15,280 | 14,437 | 14,057 | 1,603 | 10,899 | 1,555 |
| 2019-09-01 | 12,629 | 12,855 | 12,477 | 1,435 | 9,827 | 1,215 |
| 2019-10-01 | 15,350 | 15,813 | 15,338 | 1,753 | 12,013 | 1,572 |
| 2019-11-01 | 11,901 | 12,720 | 12,379 | 1,354 | 9,691 | 1,334 |
| 2019-12-01 | 12,535 | 11,649 | 11,350 | 1,235 | 8,761 | 1,354 |
| 2020-01-01 | 13,715 | 14,590 | 14,212 | 1,483 | 10,914 | 1,815 |
| 2020-02-01 | 13,171 | 13,445 | 13,167 | 1,417 | 10,277 | 1,473 |
| 2020-03-01 | 14,335 | 9,935 | 9,789 | 1,121 | 7,412 | 1,256 |
| 2020-04-01 | 10,844 | 4,563 | 4,528 | 578 | 3,344 | 606 |
| 2020-05-01 | 10,783 | 4,542 | 4,513 | 582 | 3,229 | 702 |
| 2020-06-01 | 12,653 | 9,012 | 8,950 | 1,605 | 6,293 | 1,052 |
| 2020-07-01 | 12,018 | 8,054 | 8,036 | 1,123 | 5,928 | 985 |
| 2020-08-01 | 11,568 | 8,477 | 8,458 | 1,194 | 6,199 | 1,065 |
| 2020-09-01 | 12,279 | 9,467 | 9,441 | 1,190 | 7,148 | 1,103 |
| 2020-10-01 | 13,209 | 11,801 | 11,736 | 1,526 | 9,049 | 1,161 |
| 2020-11-01 | 11,280 | 9,817 | 9,747 | 1,189 | 7,582 | 976 |
| 2020-12-01 | 12,145 | 10,243 | 10,189 | 1,235 | 7,886 | 1,068 |
| 2021-01-01 | 11,862 | 9,648 | 9,624 | 1,262 | 7,242 | 1,120 |
| 2021-02-01 | 12,003 | 11,485 | 11,444 | 1,471 | 8,755 | 1,218 |
| 2021-03-01 | 13,689 | 14,149 | 13,993 | 1,720 | 10,748 | 1,525 |
| 2021-04-01 | 12,805 | 12,400 | 12,283 | 1,390 | 9,487 | 1,406 |
| 2021-05-01 | 12,351 | 12,518 | 12,393 | 1,437 | 9,492 | 1,464 |
| 2021-06-01 | 13,608 | 14,160 | 14,021 | 1,589 | 10,783 | 1,649 |

**Table A.6:** **Quantifying racial disparities in incarcerated population declines.** Using yearly data from the Bureau of Justice Statistics [2, 18], we compare the incarcerated Black and White populations in each state from end-of-year 2019 to end-of-year 2020. The last column shows the expected additional decreases in incarcerated Black people if the Black and White incarcerated populations declined at the same rates.

| State Prisons | Incarcerated Total |  |  | Incarcerated White |  |  | Incarcerated Black |  |  | Expected additional reduction<br>in incarcerated Black population<br>under White decarceration rates |
| --- | --- | --- | --- | --- | --- | --- | --- | --- | --- | --- |
|  | 2019 | 2020 | % diff | 2019 | 2020 | % diff | 2019 | 2020 | % diff |  |
| Alabama | 28,304 | 25,328 | 10.51% | 13,454 | 11,607 | 13.73% | 14,631 | 13,519 | 7.60% | -897 |
| Alaska | 4,475 | 4,578 | -2.30% | 1,863 | 1,850 | 0.70% | 454 | 464 | -2.20% | -14 |
| Arizona | 42,441 | 37,731 | 11.10% | 16,523 | 14,613 | 11.56% | 6,233 | 5,614 | 9.93% | -102 |
| Arkansas | 17,759 | 16,094 | 9.38% | 9,767 | 8,691 | 11.02% | 7,282 | 6,721 | 7.70% | -242 |
| California | 122,687 | 97,328 | 20.67% | 25,510 | 18,819 | 26.23% | 35,056 | 28,578 | 18.48% | -2,717 |
| Colorado | 19,785 | 16,168 | 18.28% | 9,026 | 7,363 | 18.42% | 3,507 | 3,016 | 14.00% | -156 |
| Connecticut | 12,823 | 9,559 | 25.45% | 3,803 | 2,619 | 31.13% | 5,457 | 4,201 | 23.02% | -443 |
| Delaware | 5,692 | 4,710 | 17.25% | 1,932 | 1,493 | 22.72% | 3,436 | 2,934 | 14.61% | -279 |
| Florida | 96,009 | 81,027 | 15.60% | 38,335 | 31,647 | 17.45% | 45,121 | 38,665 | 14.31% | -1,416 |
| Georgia | 54,816 | 47,141 | 14% | 19,795 | 16,650 | 15.89% | 32,656 | 28,406 | 13.01% | -939 |
| Hawaii | 5,279 | 4,171 | 20.99% | 1,198 | 949 | 20.78% | 323 | 186 | 42.41% | 69 |
| Idaho | 9,437 | 8,171 | 13.42% | 7,076 | 5,966 | 15.69% | 287 | 267 | 6.97% | -26 |
| Illinois | 38,259 | 29,729 | 22.30% | 12,223 | 9,271 | 24.15% | 20,851 | 15,866 | 23.91% | -51 |
| Indiana | 27,180 | 23,944 | 11.91% | 16,878 | 14,748 | 12.62% | 8,852 | 7,888 | 10.89% | -154 |
| Iowa | 9,282 | 8,307 | 10.50% | 6,047 | 5,350 | 11.53% | 2,355 | 2,129 | 9.60% | -46 |
| Kansas | 10,177 | 8,779 | 13.74% | 5,854 | 5,030 | 14.08% | 2,755 | 2,399 | 12.92% | -32 |
| Kentucky | 23,082 | 18,552 | 19.63% | 17,542 | 13,936 | 20.56% | 4,877 | 4,052 | 16.92% | -178 |
| Louisiana | 31,609 | 26,964 | 14.70% | 10,436 | 8,710 | 16.54% | 21,061 | 18,143 | 13.85% | -566 |
| Maine | 2,185 | 1,714 | 21.56% | 1,779 | 1,389 | 21.92% | 231 | 178 | 22.94% | 2 |
| Maryland | 18,595 | 15,623 | 15.98% | 4,310 | 3,531 | 18.07% | 13,197 | 11,120 | 15.74% | -309 |
| Massachusetts | 8,205 | 6,762 | 17.59% | 3,462 | 2,805 | 18.98% | 2,306 | 1,934 | 16.13% | -66 |
| Michigan | 38,053 | 33,617 | 11.66% | 17,224 | 14,194 | 17.59% | 20,040 | 17,231 | 14.02% | -717 |
| Minnesota | 9,982 | 8,148 | 18.37% | 4,675 | 3,744 | 19.91% | 3,586 | 2,994 | 16.51% | -123 |
| Mississippi | 19,417 | 17,577 | 9.48% | 7,133 | 6,345 | 11.05% | 12,025 | 10,998 | 8.54% | -302 |
| Missouri | 26,044 | 23,062 | 11.45% | 16,286 | 14,405 | 11.55% | 9,027 | 8,003 | 11.34% | -19 |
| Montana | 4,723 | 3,927 | 16.85% | 3,359 | 2,778 | 17.30% | 131 | 103 | 21.37% | 5 |
| Nebraska | 5,682 | 5,306 | 6.62% | 2,943 | 2,733 | 7.14% | 1,559 | 1,454 | 6.74% | -7 |
| Nevada | 12,840 | 11,422 | 11.04% | 5,537 | 4,830 | 12.77% | 3,990 | 3,555 | 10.90% | -75 |
| New Hampshire | 2,691 | 2,352 | 12.60% | 3,268 | 1,978 | 39.47% | 140 | 159 | -13.57% | -75 |
| New Jersey | 18,613 | 12,830 | 31.07% | 3,978 | 2,619 | 34.16% | 11,372 | 7,772 | 31.66% | -286 |
| New Mexico | 6,723 | 5,500 | 18.19% | 1,687 | 1,477 | 12.45% | 463 | 398 | 14.04% | 7 |
| New York | 43,500 | 34,128 | 21.54% | 10,421 | 7,685 | 26.25% | 21,109 | 17,066 | 19.15% | -1,500 |
| North Carolina | 34,079 | 29,461 | 13.55% | 13,553 | 11,614 | 14.31% | 17,545 | 15,148 | 13.66% | -114 |
| North Dakota | 1,794 | 1,401 | 21.91% | 1,098 | 848 | 22.77% | 186 | 167 | 10.22% | -24 |
| Ohio | 50,338 | 45,036 | 10.53% | 25,070 | 22,470 | 10.37% | 21,752 | 19,454 | 10.56% | 42 |
| Oklahoma | 25,679 | 22,462 | 12.53% | 13,184 | 11,165 | 15.31% | 6,689 | 5,940 | 11.20% | -276 |
| Oregon | 14,961 | 12,753 | 14.76% | 10,757 | 9,257 | 13.94% | 1,436 | 1,179 | 17.90% | 56 |
| Pennsylvania | 45,702 | 39,357 | 13.88% | 20,086 | 17,131 | 14.71% | 20,846 | 18,240 | 12.50% | -461 |
| Rhode Island | 2,740 | 2,227 | 18.72% | 1,142 | 853 | 25.31% | 782 | 661 | 15.47% | -77 |
| South Carolina | 18,608 | 16,157 | 13.17% | 6,950 | 5,896 | 15.17% | 10,993 | 9,680 | 11.94% | -355 |
| South Dakota | 3,801 | 3,250 | 14.50% | 1,998 | 1,727 | 13.56% | 289 | 266 | 7.96% | -17 |
| Tennessee | 26,349 | 22,685 | 13.91% | 14,633 | 12,359 | 15.54% | 11,012 | 9,656 | 12.31% | -356 |
| Texas | 158,429 | 135,906 | 14.22% | 53,597 | 45,596 | 14.93% | 51,584 | 44,760 | 13.23% | -877 |
| Utah | 6,665 | 5,446 | 18.29% | 4,048 | 3,245 | 19.84% | 471 | 400 | 15.07% | -23 |
| Vermont | 1,608 | 1,284 | 20.15% | 1,386 | 1,108 | 20.06% | 141 | 123 | 12.77% | -11 |
| Virginia | 36,091 | 31,838 | 11.78% | 15,005 | 13,266 | 11.59% | 19,808 | 17,414 | 12.09% | 98 |
| Washington | 19,261 | 15,724 | 18.36% | 11,262 | 8,843 | 21.48% | 3,275 | 2,777 | 15.21% | -206 |
| West Virginia | 6,800 | 6,044 | 11.12% | 5,819 | 5,144 | 11.60% | 875 | 802 | 8.34% | -29 |
| Wisconsin | 23,956 | 20,298 | 15.27% | 10,815 | 8,879 | 17.90% | 10,003 | 8,542 | 14.61% | -330 |
| Wyoming | 2,479 | 2,087 | 15.81% | 1,864 | 1,591 | 14.65% | 129 | 89 | 31.01% | 21 |
| Nationwide | 1,255,689 | 1,063,665 | 15.29% | 515,591 | 430,817 | 16.44% | 492,186 | 421,311 | 14.40% | -14,593 |

**Table A.7:** **States report race differently.** In this table, the races listed (and their ordering) are copied exactly as reported by the state. All states include White or Caucasian as a race, most include Black or African American, many include Latino or Hispanic. Michigan reports White and Non-White as its race categories.

| State | Races included in incarcerated population reports |
| --- | --- |
| Alabama | Black, White, Other |
| Alaska | Alaska Native, Asian/Pacific Islander, Black, Caucasian, Hispanic/Latino, Other/Unknown |
| Arizona | Hispanic, Caucasian, African-American, Native American, Other |
| Arkansas | Asian, Black, Hispanic, Native American Indian, Native Hawaiian or Pacific Islander, White, Other, Unknown |
| California | Hispanic, Black, White, Other |
| Colorado | Caucasian, Hispanic, African American, Native American, Asian |
| Connecticut | White, Black, Hispanic, American Indian, Asian |
| Delaware | Black, White, Asian or Pacific Islander, American Indian/Alaskan, Unknown |
| District of Columbia | Black, White, Hispanic, Other |
| Florida | White, Hispanic, Black, Other |
| Georgia | White, Black, Other, Asian, Unknown, Hispanic, Native American |
| Hawaii | American Indian, African American, Caucasian, Chinese, Filipino, Guam/Pacific Islander, Native Hawaiian, Hispanic, Japanese, Korean, Samoan, Other, Unknown |
| Idaho | Asian, Black or African American, Hispanic or Latino, Indian, Other, Unknown, White |
| Illinois | American Indian or Alaskan Native, Asian, African American or Black, Hispanic or Latino, Native Hawaiian or other Pacific Islander, White, Other, Two or More Races |
| Indiana | White, Black, Hispanic, Undetermined, American Indian/Alaskan Native, Asian/Pacific Islander |
| Iowa | White, Black, Hispanic, American Indian or Alaskan Native, Asian or Pacific Islander, Unknown |
| Kansas | White, Black, American Indian, Asian; reports Hispanic and Non Hispanic separately as ethnicity |
| Kentucky | White, Black, Hispanic, Bi-Racial, Asian, Native American, Other |
| Louisiana | White, Black, Other |
| Maine | Asian, Black or African American, Native American, Native Hawaiian, Two or More Races, Unknown, White |
| Maryland | Black, White, Latinx, Indian, Asian, Unknown |
| Massachusetts | Caucasian, African American, Hispanic, Asian, Other, Native American, Native Hawaiian/Pacific |
| Michigan | White, Non-White |
| Minnesota | White, Black, American Indian, Asian, Unknown/Other; reports Hispanic separately as ethnicity |
| Mississippi | Black, White, Hispanic, Native American, Asian, Data Unavailable |
| Missouri | Asian, Black, Hispanic, Native American, Unknown, White |
| Montana | American Indian/Alaska Native, Black, White, Asian/Pacific Islander |
| Nebraska | White, Black, Hispanic/Latino, American/Alaskan Native, Asian, Hawaiian/Pacific Islander, Other, Data Unavailable |
| Nevada | African American, American Indian, Asian, Caucasian, Hispanic, Other |
| New Hampshire | White, Black, Native American, Asian, Other/Unreported |
| New Jersey | Black, White, Hispanic, Asian, American Indian, Other or Not Coded |
| New Mexico | Asian, Black/African American, Hispanic Black, Hispanic Native Indian, Hispanic White, Native American/Alaskan, Native American, Pacific Islander, Unknown, White |
| New York | White, African American, Hispanic, Native American, Asian, Other, Unknown |
| North Carolina | White, Black/African American, American Indian/Alaskan Native, Asian/Asian American, Other, Unknown |
| North Dakota | Native American, Black, Hispanic, White, Other |
| Ohio | White, Black, Other Race |
| Oklahoma | African American, Caucasian, Hispanic, Native American, Other |
| Oregon | Asian, Black, Hispanic, American Indian, White, Unknown, Pacific Islander |
| Pennsylvania | Black, White, Hispanic, Other |
| Rhode Island | Black, White, Asian, Hispanic, Other |
| South Carolina | Black, White, Other |
| South Dakota | White, Native American, African American, Hispanic, Asian, Native Hawaiian/Pacific Islander |
| Tennessee | White, Black, Other |
| Texas | Black, White, Hispanic, Other and Unknown Race |
| Utah | Asian/Pacific Islander, Black, Hispanic, Native American/Alaskan, White, Unknown, White |
| Vermont | American Indian/Alaskan Native, Asian/Pacific Islander, Black or African American, White, Some Other Race/Unknown; reports Hispanic/Latinx/Spanish Identification separately as ethnicity |
| Virginia | Black, White, Other |
| Washington | White, Black, American Indian/Alaska Native, Asian/Pacific Islander, Other, Unknown |
| West Virginia | American Indian/Alaska Native, Asian, Black, Hispanic or Latino, Multi-Racial or Other, Native Hawaiian or Pacific Islander, White |
| Wisconsin | White, Black, American Indian/Alaskan Native, Asian or Pacific Islander, Unknown |
| Wyoming | Caucasian, African American, Hispanic, Native American, Other Race |

## Footnotes

1 Table A.4 is still awaiting answers from Washington D.C., Alabama, and Louisiana.

2 Note: OSCA is the organization that compiled the data and are not the custodians of the data. Clerks of the court record each defendant’s case data via the Offender Based Transaction System. Any conclusions or analysis that will derive from this dataset are solely those of the individual author(s) or the person(s) who did the analysis and not of the Florida Office of State Court Administrator.

## Notes

### Competing Interest Statement

The authors have declared no competing interest.

### Funding Statement

This study did not receive any funding.

### Summary of Updates

Updated main text, restructured manuscript and supplemental information.

